# Panomics reveals patient-individuality as the major driver for colorectal cancer progression

**DOI:** 10.1101/2022.10.06.22280355

**Authors:** Friederike Praus, Axel Künstner, Thorben Sauer, Michael Kohl, Katharina Kern, Steffen Deichmann, Ákos Végvári, Tobias Keck, Hauke Busch, Jens K Habermann, Timo Gemoll

## Abstract

Colorectal cancer (CRC) is one of the most prevalent cancers, with over one million new cases. The prognosis of CRC considerably depends on the disease stage and metastatic status. As precision oncology for patients with CRC continues to improve, this study aims to integrate genomic, transcriptomic, and proteomic analyses to identify significant expression differences during colorectal progression using a unique set of paired patient samples concerning tumor heterogeneity.

We analyzed fresh-frozen tissue samples of matched healthy colon mucosa, colorectal carcinoma, and liver metastasis from same patients prepared under strict cryogenic conditions. While somatic mutations of known cancer-related genes were analyzed using Illumina’s TruSeq Amplicon Cancer Panel, the transcriptome was assessed comprehensively using Clariom D microarrays. The global proteome was evaluated by liquid chromatography-coupled mass spectrometry (LC-MS/MS) and validated by two-dimensional difference in-gel electrophoresis. Subsequent unsupervised principal component clustering, statistical comparisons, and gene set enrichment analyses were calculated using differential expression results.

While panomics revealed low RNA and protein expression of CA1, CLCA1, MATN2, AHCYL2, and FCGBP in malignant tissues compared to healthy colon mucosa, no differentially expressed RNA or protein targets were detected between tumor and metastatic tissues. Subsequent intra-patient comparisons revealed highly specific expression differences (e.g., SRSF3, OLFM4, and CEACAM5) associated with a patient-individual transcriptome and proteome.

In conclusion, the results highlight the importance of inter- and intra-tumor heterogeneity alongside the individual, patient-paired evaluation for clinical studies. Next to changes among groups reflecting colorectal cancer progression, we identified significant expression differences between patient-individual normal colon mucosa, primary tumor, and liver metastasis, which could speed up the implementation of precision oncology in the future.

## Introduction

Colorectal cancer (CRC) is the third most common cancer and cause of cancer-related death [1]. Whereas early-stage CRC has a relatively good prognosis with a 5-year survival of almost 90 %, metastatic CRC (mCRC) prognosis is poor with a 5-year survival of only 15 % [2]. In this context, over 50 % of CRC patients either show liver metastasis at the primary diagnosis or develop progress shortly afterward [3, 4]. However, knowledge about disease progression and how tumor heterogeneity contributes to metastasis or treatment tolerance is still limited [5].

To understand the underlying molecular changes that drive the carcinogenesis of mCRC, the Cancer Genome Atlas (TCGA) has conducted extensive genomic, epigenomic, and transcriptomic profiling studies to identify distinguishing features of mCRC [6-9]. These studies have underscored the value of molecular characterization in addition to histological assessment for the stratification of mCRC patients while identifying genomic features unique to the tumor genesis of mCRC [10]. Although it has been shown that the mutation status of specific oncogenes such as *HER2* in breast cancers, *EGFR* in lung cancers, and *KRAS* in colon cancers are crucial for the targeted treatment [11, 12], genomic markers fail to find eligible patients and to predict therapy outcomes in the majority of cases. Marquart and colleagues estimated in 2018 that for patients with advanced or metastatic cancer, only 8.3 % would be eligible for genome-driven drugs, and only 4.9 % would benefit from these drugs [13]. Therefore, combining proteomics and genomics to proteogenomic analyses can improve individualized cancer medicine [14].

The proteome with this represents a central position. While it is regulated downstream by the genetic expression and reacts to signals and treatments of the environment, the main task of proteins is to mediate the biochemical activities of cells and organs [15]. As such, proteins are perfectly eligible to determine the significance of a physiological phenotype and a point of intervention for drug and health treatments [16-18]. Several preclinical studies have identified the proteome of CRCs to underline the biological changes affecting CRC. Interestingly, most of these proteomic studies have focused on differentially expressed proteins, wherein the patients of the primary colorectal tumor sample and the liver metastasis sample differed. For instance, Li et al. identified metastasis-related factors in 2020 that were differentially expressed in synchronous solitary liver metastasis compared to primary colon cancer [19]. By integrating the genomics, proteomics, and phosphoproteomics of 480 clinical tissues, molecular signatures characterized three CRC subtypes. The authors detected high similarities with primary tumors on genetic but not proteomic levels. In line, Sardo et al. recently reviewed panomics approaches in CRC beyond genomic data and presented, e.g., predictive proteomic targets in clinical settings [20]. However, little is currently known about panomics applied to individual-matched patient samples.

Characterizing the link between intra- and inter-tumor heterogeneity, we present a global analysis of genomics, transcriptomics, and proteomics in mCRC from paired clinical samples.

## Material and Methods

### Overview of the patient cohort

Paired tissue samples from normal adjacent colon mucosa (NM), corresponding primary colorectal carcinomas (T), and corresponding liver metastases (LM) were obtained from four patients (P1-4). For one patient, we collected eight samples: normal colon mucosa, six samples from different locations within the primary tumor, and one sample from the corresponding liver metastasis, respectively. **Table 1** present all patient characteristics. All patients were diagnosed with metastasized colorectal carcinoma and received a primary resection. Samples were surgically removed at the Department of Surgery, University Hospital Schleswig-Holstein, Campus Lübeck, and stored in liquid nitrogen until further processing. The Ethics Committee of the University of Lübeck gave ethical approval for this work (No. 07-124 and 16-282).

**Table 1:**
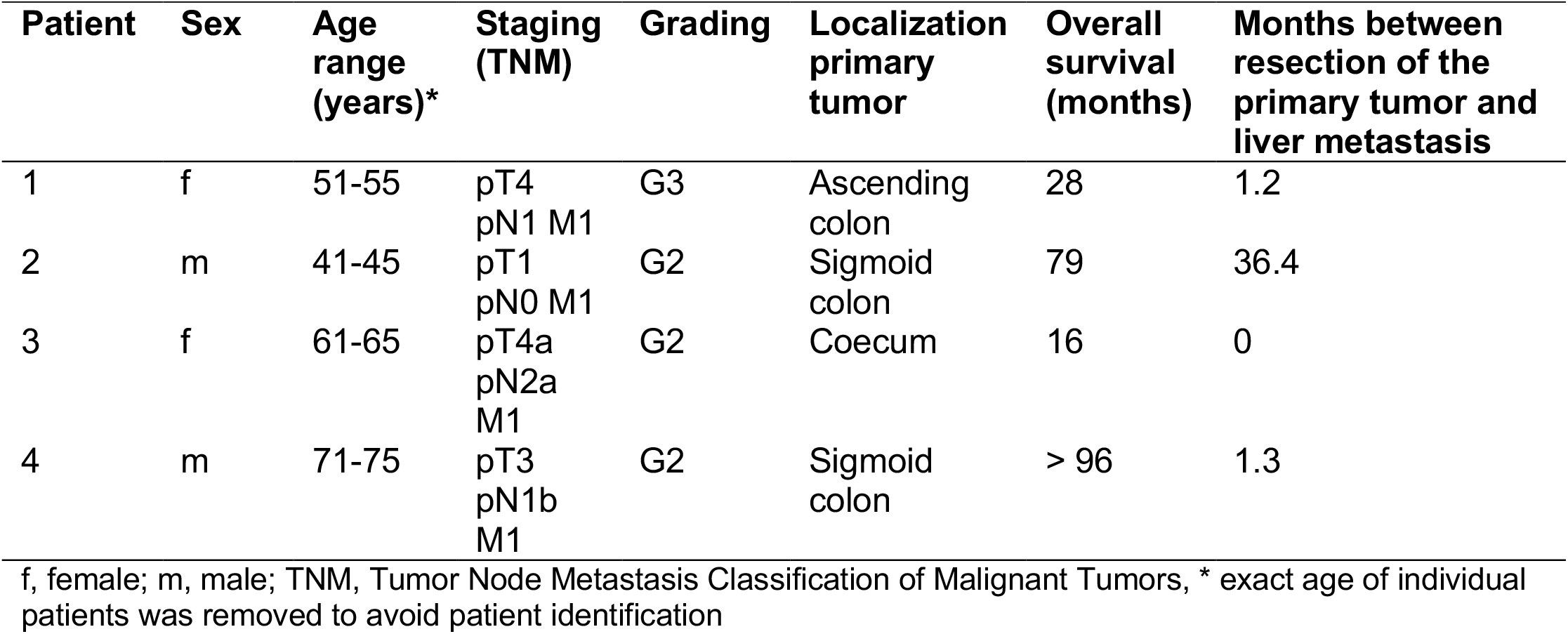
Clinical parameters of the patient cohort.

| Patient | Sex | Age range (years)* | Staging (TNM) | Grading | Localization primary tumor | Overall survival (months) | Months between resection of the primary tumor and liver metastasis |
| --- | --- | --- | --- | --- | --- | --- | --- |
| 1 | f | 51-55 | pT4<br>pN1 M1 | G3 | Ascending colon | 28 | 1.2 |
| 2 | m | 41-45 | pT1<br>pN0 M1 | G2 | Sigmoid colon | 79 | 36.4 |
| 3 | f | 61-65 | pT4a<br>pN2a<br>M1 | G2 | Coecum | 16 | 0 |
| 4 | m | 71-75 | pT3<br>pN1b<br>M1 | G2 | Sigmoid colon | > 96 | 1.3 |

f, female; m, male; TNM, Tumor Node Metastasis Classification of Malignant Tumors, \* exact age of individual patients was removed to avoid patient identification

### Sample preparation concerning intra-tumor heterogeneity

To allow for subsequent downstream analysis of intra-tumor heterogeneity while maintaining low temperatures, all samples were manually divided into halves on dry-ice pre-chilled plates. 6 µm sections from both sides of the two fractions were cut and stained with hematoxylin and eosin (H&E) for histopathologic classification. Semi-automated frozen aliquots (1.5 mm coring) were subsequently taken from the areas with the highest tumor cell (or mucosal cell) representativity by using the CryoXtract CXT350 at a temperature below -100 °C and transferred to 700 µl lysis buffer (Qiagen buffer RLT plus, 1 % β-mercaptoethanol). Supplemental data present representative images before and after the coring process (**Supplemental Figure 1**).

For the extraction of nucleic acids and proteins, the AllPrep^®^ DNA/RNA Micro Kit (Qiagen, Germany) was used according to manufacturers’ instructions with additional steps from the AllPrep^®^ DNA/RNA/Protein Mini Kit (Qiagen, Germany) and the PureLink™ DNase (Invitrogen, USA). This extraction protocol resulted in one 100 µl DNA, 60 µl RNA, and 10 µl miRNA solution as well as in a protein pellet dissolved in 200 µl lysis buffer. 100 µl of each protein sample was purified with the ReadyPrep 2-D Cleanup Kit (Bio-Rad, USA). The purified protein pellet was dissolved in 22 µl DIGE buffer [30 mM TRIS, 7 M urea, 2 M thiourea, 4 % (w/v) CHAPS]. A 2 µl aliquot was used to determine total protein concentration with the EZQ™ Protein Quantitation Kit (Life Technologies, USA). The remaining protein solution was stored at -80 °C until further analysis.

### Targeted detection of somatic mutations in cancer-related genes

The TruSeq™ Amplicon Cancer Panel (TSACP, Illumina, San Diego, CA, USA) is a multiplexed targeted amplicon sequencing (tNGS) assay aiming to detect somatic mutations frequently reported as related to cancer. The library preparation was carried out according to Illumina’s TSACP standard protocol. All samples were sequenced in two runs with the MiSeq™ next-generation sequencing (NGS) system. To this end, the MiSeq Reagent Kit V2 was utilized at 300 cycles.

Several analysis procedures, including demultiplexing and FASTQ file generation, were performed using the MiSeq reporter software (Real-Time-Analysis (RTA), Version: 1.18.54). Starting with the FASTQ files containing raw paired-end data, an in-house software pipeline was applied for data analysis. Briefly, the reads were mapped to the reference genome (GRCH38/hg38) with the Burrows-Wheeler Aligner (BWA-MEM v.0.7.17-4). PICARD TOOLS (v.2.3.2-1) were used to sort the resulting SAM files and for conversion in the BAM format. Adjustment of quality scores was carried out using the Base Quality Score Recalibration (BQSR) method provided by GATK (v. 4.2.6.1). The preprocessed BAM files are input for several working steps that are provided by tools of the GATK best practices workflow (GATK v. 4.2.6.1) for the detection of somatic short variants (single nucleotide variants, SNVs, and insertions/deletions, Indels). This workflow includes the usage of Mutect2 for the computation of a basic callset of candidate variants and subsequent filtering with the FilterMutectCalls algorithm of GATK. Ensembl Variant Effect Predictor (VEP, release 106) was used to annotate the filtered variants with information regarding the effects of the detected somatic variants. Next, annotations (in mutation annotation format, MAF) were imported into R (v. 4.2.0). Several cleaning/filtering procedures were applied (e.g., entries in the call set that are not mapped to genes included in the TSCAP, variants with low coverage or variants with population allele frequency >0.001 in the gnomAD or 1k genome databases were removed). The filtered data was set as input for the R package maftools (package version 2.12.0) for data visualization and the computation of summary metrics for the data set [21].

### Transcriptome profiling

Data generation was performed according to the vendor’s original protocol using the GeneChip™ WT Pico Reagent Kit (Thermo Scientific, Waltham, Massachusetts, USA), followed by hybridization on the Clariom D array (Clariom™ D Pico Assay, human, Thermo Scientific). Clariom D array data were imported in R (v4.1.2) using the oligo package (v1.60.0) preprocessed utilizing the robust multichip average algorithm (RMA). Annotations were added using the *annotateEset* function as implemented in the affycoretools (v1.68.0) package and clariomdhumantranscriptcluster.db (v8.8.0) as annotations. Probes not matching known genes were removed before analysis.

### Mass spectrometry profiling

Equal aliquots of 47 µg of protein from each protein sample were diluted with DIGE buffer [30 mM TRIS, 7M urea, 2M thiourea, 4 % (w/v) CHAPS] to a total volume of 50 µl and cleaned with a filter-aided sample preparation protocol (FASP) [22]. Dried protein pellets were diluted in 40 µl of 5 % formic acid before mass spectrometric analysis, and half of the sample was prepared on a StageTip as previously described [23]. Peptides were separated chromatographically using a 25 cm long C18 column (SilicaTip™ 360 µm OD, 100 µm ID, New Objective, USA) in an EASY-nLC1000™ nanoflow LC system (Thermo Fisher Scientific, USA). With a 300 nL/min flow rate, peptides were eluted at a linear gradient from 2 to 26 % solvent B (0.1 % formic acid in 98 % acetonitrile) for 120 min. Mass spectrometric detection of eluted peptides was carried out using a Q Exactive™ Plus hybrid quadrupole-Orbitrap™ mass spectrometer (Thermo Fisher Scientific, Bremen, Germany) in the data-dependent mode. The survey mass spectrum was acquired at the resolution of 140,000 (at *m/z* 200) in the range of *m/z* 300-1650, targeting 5·10^6^ ions. The MS/MS data for the 16 most intense precursors were obtained with a higher-energy collisional dissociation (HCD) set at 28 % normalized collision energy following isolation of precursor ions with 4 Th targeting 2·10^5^ ions with charge z>1 at a resolution of 17,500.

Tandem mass spectra were extracted using Raw2MGF (in-house-written program), and the resulting Mascot generic files (.mgf) were searched against a concatenated SwissProt protein database (Human taxonomy) using Mascot 2.3.0 search engine (Matrix Science Ltd., London, UK). Carbamidomethylations of cysteines were set as a fixed modification and deamidation of asparagine and glutamine as well as oxidation of methionine were set as variable modifications. Up to two missed tryptic cleavages were allowed and the mass tolerance was set to 10 ppm and 0.05 Da for the precursor and fragment ions, respectively. Only peptides with individual MS/MS Mascot scores above the significant threshold corresponding to E<0.05 were accepted. Only proteins identified with at least two peptides with a significant score and a 0.25 % false discovery rate (FDR) were considered for further quantification.

Acquired mass spectra were analyzed with in-house developed Quanti software (v2.5.4.4) using the relative abundance of proteins identified with more than two unique peptides [24]. Minimal requirements were a peptide length of six amino acids and a false discovery rate of 0.01. The areas of the chromatographic peaks were taken as the peptide abundances and the same peptides were quantified in each nano-LC-MS/MS data file using accurate mass and the order of elution as identifiers. Following settings were applied: (1) enzyme “trypsin”, (2) fixed modifications “cysteine carbamidomethyl”, (3) optional modifications “methionine oxidation, asparagine, and glutamine deamidation, N-terminal acetylation” and (4) a maximum of two missed cleavages. Results were analyzed in the R scripting and statistical environment. Data were normalized by calculating the summed intensities of all proteins in each sample and the median of all these summed intensities over the entire sample set. Each quantitative value was multiplied by the median/summed intensity and the resulting values were log2 transformed. Differences in relative protein abundances between treated and control samples were assessed by moderated t-test using the limma package [25]. Furthermore, Benjamini-Hochberg correction for multiple comparisons was applied.

### Protein profiling by two-dimensional gel electrophoresis

Paired patient clustering was validated by multiplex fluorescent two-dimensional gel-electrophoresis (2-D DIGE). The Refraction-2D™ Labelling Kit (NH DyeAGNOSTICS, Germany) was used to label 50 µg of each protein sample, a pool of the tumor samples from P3, and an internal standard described previously [26]. Briefly, proteins were applied to an immobilized pH gradient gel strip (pH range 4-7) for active rehydration and separated by SDS-Page on precast 12.5 % acrylamide gels. Gel images were acquired with a Typhoon FLA 9000 scanner (GE Healthcare, UK) and analyzed with Progenesis SameSpots (Nonlinear Dynamics, USA; v4.5). Spots were aligned to a reference image, automatically detected, manually corrected, and normalized against the internal standard.

### Statistical analysis

The LC-MS/MS data were analyzed using the R statistical environment. Proteins with missing values in at least one sample and proteins with unknown gene names were removed. The remaining data were normalized using quantile normalization from the preprocessCore R package (v1.52.1).

To cluster samples based on RNA and protein expression profiles, principal component analysis was performed using the 100 proteins with the highest variance (FactoMineR R package, v2.4, [27]). Differentially abundant genes and proteins were detected using a linear model approach from the limma package for R (v3.46.0) [25]. The two-group comparisons NM vs. T, NM vs. LM, and T vs. LM were carried out across all patients. The within-patient correlation was estimated using the duplicateCorrelation function Field [28] to correct repeated measurements in the same patients. A significance level of q <0.01 and a relevant fold change (FC) of |log_2_FC| >1 were applied for two-group comparisons. Individual patient comparisons were evaluated by calculating the correlation factor ρ and plotting protein FCs of NM vs. T and NM vs. LM against each other. Due to the more significant difference between individual patient comparisons, the effect size was set to |log_2_FC| >2. Identified proteins of each sample were further enriched against the HALLMARK gene sets (MSigDB v7.1) using Generally Applicable Gene Set Enrichment (GAGE) as implemented in the R package gage (v2.40.1) [28, 29].

2-D DIGE was analyzed by SameSpots software (v4.5, Nonlinear Dynamics, USA). For ANOVA two-group comparisons, a p-value <0.05 and a |log_2_FC| >1 were considered significant.

### Data availability

Transcriptomic expression data are available at Gene Expression Omnibus (GEO, https://www.ncbi.nlm.nih.gov/geo/) with the accession number GSE206800. The mass spectrometry proteomics data are deposited to the ProteomeXchange Consortium via the PRIDE partner [30] repository with the data set identifier PXD036434.

## Results

To compare the difference between fresh frozen samples derived from patients with advanced-stage colorectal cancer, we performed panomics by TruSeq™ Amplicon sequencing, Clariom D arrays, quantitative mass spectrometric profiling, and 2-dimensional gel electrophoresis. For all patients, primary tumor tissue was taken before chemotherapy. All patients had synchronous liver metastasis at the time of diagnosis (**Table 1**). Except for one patient (P2), all metastatic tissues were taken before exposure to chemotherapy. For patient 4, screening six different primary tumor locations was also possible.

### Characterization of paired patient samples using gene expression data and mutation analysis

To identify genomic features, we analyzed mutational data of the selected cohort (targeted sequencing). Frequently mutated genes were *APC* (100 %), *GNA11* (100 %), *TP53* (100 %), *ERBB2* (75 %), *KRAS* (62 %), and *ATM* (62 %). All 24 genes with a detectable mutation are presented in **Supplemental Figure 2**. We did not observe any significantly unbalanced distribution between malignant groups and individual patients, considering the mutated genes.

Gene expression profiles of 25,161 genes were retrieved from microarrays and visualized using the top 2,000 most variable genes (**Figure 1a**). Remarkably, patients and not disease groups cluster together. In total, 130 genes were identified as significantly differentially expressed between normal adjacent colon mucosa (NM) and corresponding primary colorectal carcinomas (T) samples (q-value < 0.01; |log_2_ FC| > 2; 62 higher expressed in NM, 68 higher expressed in T; **Figure 1b**) and 154 genes between NM and corresponding liver metastases (LM) (81 higher in NM, 73 higher in LM, **Figure 1c**). Gene expressions showed no differences between LM and T (**Figure 1d**).

**Figure 1:**
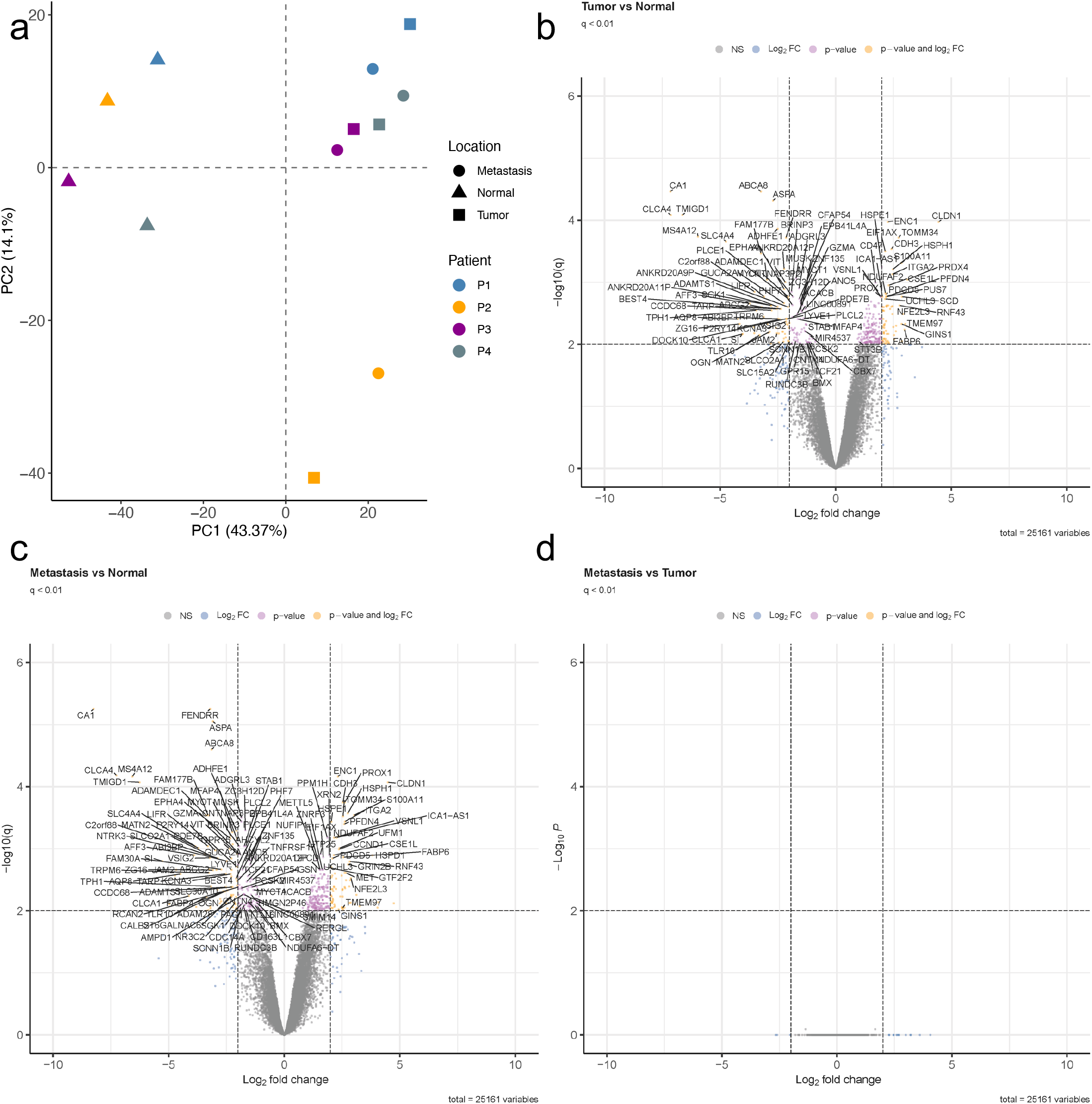
Unsupervised principal component analysis (a) and volcano plots of differentially expressed genes between all three group comparisons of NM, T, and LM (b-d). PCA plot displays all four individual patients (P1, blue; P2, yellow; P3, purple; P4, grey). X- and y-axes show the first and second principal components, respectively. Volcano plots are presented with the fold-change of the corresponding comparison in logarithmic scale (x-axis) against the q-value (y-axis) of the (b) tumor vs. normal mucosa, (c) metastasis vs. normal mucosa, and (d) metastasis vs. tumor comparison. Significance thresholds (q-value <0.01 and |log_2_FC| threshold of >2) are indicated by dashed lines. Genes passing these cut-offs are considered significant and colored in yellow. Genes passing the q-value but not the log_2_FC threshold are colored in rose. Genes that were not significant but passed the log_2_FC threshold are indicated in blue.

### Proteomic characterization of paired patient samples

Using a robust label-free workflow, all samples indicated a high proteome depth and were included in subsequent analysis (**Supplemental Figure 3**).

A total of 2,885 proteins were identified using LC-MS/MS. After the reduction of missing values, unsupervised clustering using PCA analysis of 2,686 protein groups demonstrated one close cluster of all NM samples (**Figure 2a**). In line with the transcriptomics data, patients (P1-P4) and not disease groups (T, LM) present distinct similarities pointing to individual clinical phenotypes. The comparison between NM and T samples yielded 71 significantly differentially expressed proteins (q-value < 0.01 and |log_2_ FC| > 1), with 68 proteins (96 %) up-regulated and three proteins (4 %) down-regulated in NM samples (**Figure 2b**). Similarly, in the NM and LM comparison, 69 proteins were significantly differentially expressed (q-value < 0.01 and |log_2_ FC| > 1). Of these 69 proteins, 61 (88 %) were up-regulated, and eight (12 %) were down-regulated in NM (**Figure 2c**). The chloride channel accessory 1 protein (CLCA1) was detected with an exceptionally significant low concentration in both T and LM compared to NM samples (log_2_FC_TvsNM_ = -21.010 and log_2_FC_LMvsNM_ = -20.888).

**Figure 2:**
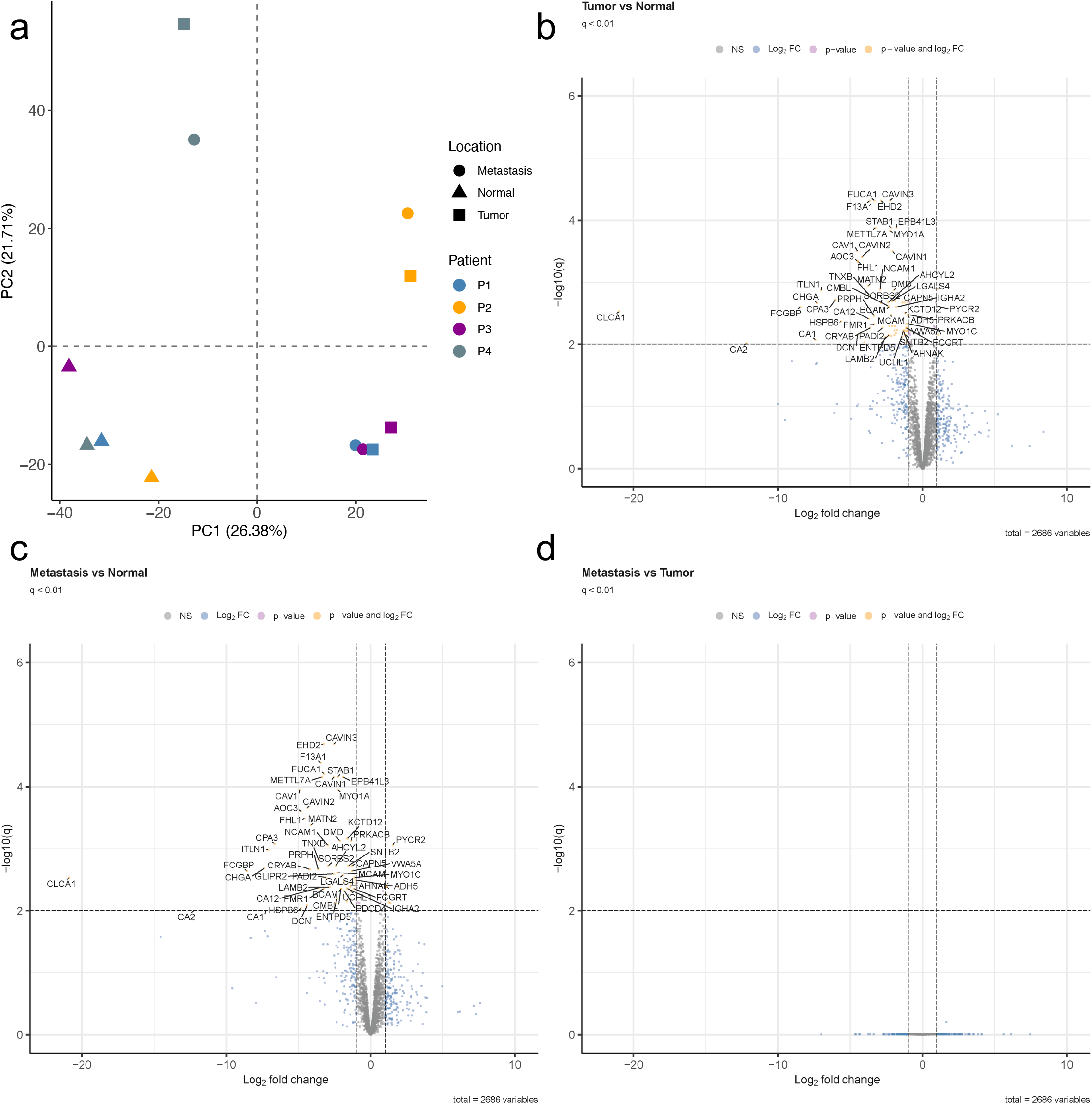
Unsupervised principal component analysis (a) and volcano plots of differentially expressed proteins between all three group comparisons of NM, T, and LM (b-d). PCA plot displays all four individual patients (P1, blue; P2, yellow; P3, purple; P4, grey). X- and y-axes show the first and second principal components, respectively. Volcano plots are presented with the fold-change of the corresponding comparison in logarithmic scale (x-axis) against the q-value (y-axis) of the (b) tumor vs. normal mucosa, (c) metastasis vs. normal mucosa, and (d) metastasis vs. tumor comparison. Significance thresholds (q-value <0.01 and |log_2_FC| threshold of >2) are indicated by dashed lines. Proteins passing these cut-offs are considered significant and colored in yellow. Proteins passing the q-value but not the log_2_FC threshold are colored in rose. Proteins that were not significant but passed the log_2_FC cutoff are indicated in blue.

The most surprising aspect of the data is that no protein could be detected to be significantly expressed between T and LM, highlighting the inter-patient heterogeneity (**Figure 2d**).

### Comparison of gene and protein expression data & validation of the global proteome by 2-D DIGE

Closer inspection of the gene (n = 25,161) and protein (n = 2,686) expression data showed an overlap of three genes/proteins for the NM vs. T comparison (CA1, CLCA1, MATN2) and five for the NM vs. LM comparison (AHCYL2, CA1, CLCA1, FCGBP, MATN2) being significantly expressed between groups. All genes/proteins were characterized by higher levels in normal material plotted based on their gene and protein abundances (**Figure 3**). In line, high expression of all targets was associated with a good prognosis in CRC using data of the Human Protein Atlas data [31, 32]. To validate the clustering results of the proteome profiling, two-dimensional gel electrophoresis was performed. SameSpots software detected 1,334 spots per gel. A PCA based on these resulted in a qualitatively similar result, indicating a clear distinction between patients and sample groups (**Supplemental Figure 4**).

**Figure 3:**
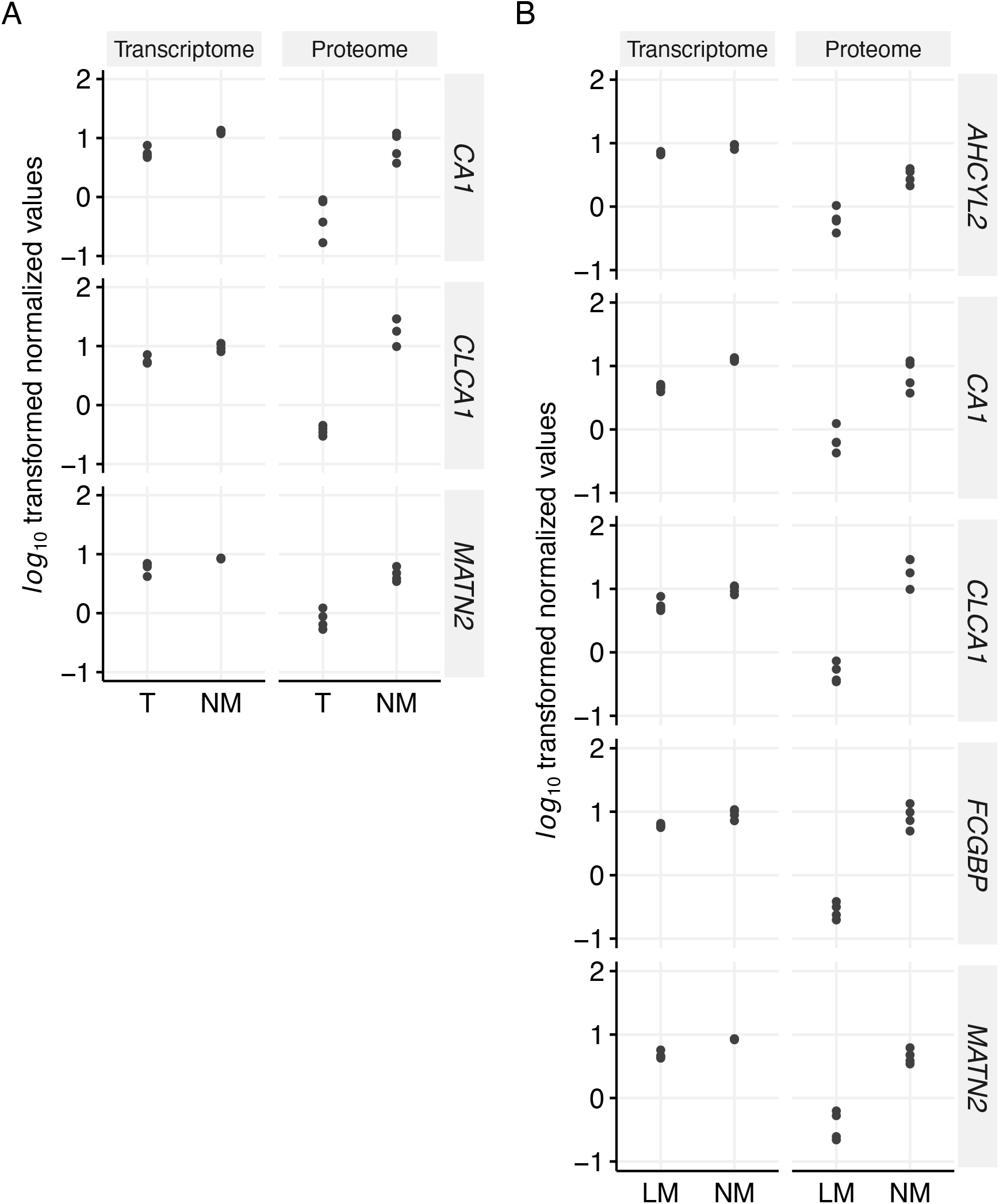
Overlap of the transcriptome and proteome data showing significant expressed targets for the NM vs. T (a) and the NM vs. LM (b) group comparisons. NM, normal mucosa; T, tumor; LM, liver metastasis

### Individual progression proteomic landscape in individual patients

Based on the group comparisons’ results, each primary tumor’s transcriptomes and proteomes were compared to their paired adjacent mucosa and associated liver metastasis. All tumor samples correlated phenotypically closely with their corresponding liver metastases but presented different gene/protein profiles compared to their adjacent normal mucosa. The Pearson correlation for gene expression and protein abundance between primary tumors and paired liver metastasis compared to their adjacent mucosa was 0.6854<r<0.8281 for the proteome and 0.6114<r<0.8133 for the transcriptome, respectively (**Supplemental Figures 5-8**).

### Patient 1: Female, right-sided CRC (pT4)

Proteome analysis of NM1, T1, and LM1 revealed proteome expression changes between initial diagnosis and metastasis surgery after one month. Of the 2,686 evaluable proteins, 294 proteins demonstrated a |log_2_ FC| > 2 in the NM1 vs. T1 and 251 in the NM1 vs. LM1 comparison (**Supplemental Figure 5a**). The comparison between T1 and LM1 presented 55 contrasting proteins (**Supplemental Figure 5b**). **Supplemental Table 1** lists the top differentially expressed proteins of the three two-group comparisons. Among proteins with a higher concentration in LM1 than T1, CD74 (log_2_ FC = 2.179) was reported as an oncogene (Network of Cancer Genes 7.0, http://ncg.kcl.ac.uk), promoting tumor growth and metastasis in various cancer types. TNC (Tenascin C) the second detectable oncogene was higher expressed in T1 (log_2_ FC = 6.522) and LM1 (log_2_ FC = 2.930) compared to NM1 but showed lower levels in LM1 compared to T1 (log_2_ FC = -3.592).

On the RNA level, 30 transcripts in the NM1 vs. T1 and 34 in the NM1 vs. LM1 comparison were differentially expressed with a |log_2_ FC| >1, respectively (**Supplemental Figure 5c**). Contrasting RNAs were presented in the T1 vs. LM1 comparison, with no genes showing a |log_2_ FC| >1 (**Supplemental Figure 5d**). **Supplemental Table 2** lists the top differentially expressed RNAs with either a higher or lower gene expression for each two-group comparison. Among RNAs with a higher concentration in LM1 compared to T1, SFRP4 was previously reported as an oncogene (Network of Cancer Genes 7.0).

The overlap of differentially expressed genes and proteins showed three molecules for the NM1 vs. T1 comparison (CA1, CLCA1, ZG16) and six for the NM1 vs. LM1 comparison (CA1, CLCA1, FABP4, MUC2, PIGR, ZG16). The concentration of CEACAM5, which has been widely applied in the clinical detection of liver metastasis from colorectal cancers, showed almost no differential expression between NM1, T1, and LM1 on the RNA and protein levels (**Supplemental Figure 5**).

### Patient 2: Male, left-sided CRC (pT1)

Proteome analysis of the second patient samples (NM2/T2 and LM2) presented proteome expression changes between initial diagnosis and metastasis surgery after 36 months. The patient received several chemotherapies during that timeframe. In total, 183 proteins in the NM2 vs. T2 and 265 proteins in the NM2 vs. LM2 comparison were differentially expressed (|log_2_ FC| > 2, **Supplemental Figures 6a & b**). The comparison between T2 and LM2 presented only 70 contrasting proteins. **Supplemental Table 3** lists the top differentially expressed proteins with either a higher or lower protein concentration for each comparison. Among proteins with a higher concentration in LM2 compared to T2, SRSF3 (Serine and Arginine Rich Splicing Factor, log_2_ FC = 2.013) was the only reported oncogene (Network of Cancer Genes 7.0). Interestingly, SRSF3 was also strongly higher expressed in T2 (log_2_ FC = 2.944) and LM2 (log_2_ FC = 4.956) compared to NM2. CEACAM5, a potential marker for colorectal progression, was strongly upregulated in T2 and LM2 compared to NM2. Regarding gene expression data, 53 RNAs revealed a |log_2_ FC| > 1 in the NM2 vs. T2 and 59 in the NM2 vs. LM2 comparison. The comparison of T2 vs. LM2 revealed 13 differentially expressed RNAs (**Supplemental Figures 6c & d**). The top differentially expressed RNAs of the three two-group comparisons are presented in **Supplemental Table 4**. The overlap of differentially expressed genes and proteins showed nine features for the NM2 vs. T2 comparison (CA1, CLCA1, CPA3, EPB41L3, JCHAIN, MATN2, MUC2, OGN, SULT1B1) and eight for the NM2 vs. LM2 comparison (CA1, CLCA1, MATN2, MEP1A, MUC2, PIGR, SULT1B1, ZG16). PIGR is an annotated healthy driver (Network of Cancer Genes 7.0) that showed lower concentrations of RNA (log_2_ FC = -1.165) and protein (log_2_ FC = -2.676) in LM2 compared to NM2.

### Patient 3: Male, left-sided CRC (pT3)

Proteome analysis of the third patient samples (NM3/T3 and LM3) detected proteome expression changes between initial diagnosis and metastasis surgery after one month. In total, 328 proteins in the NM3 vs. T3 and 264 proteins in the NM3 vs. LM3 comparison were differentially expressed (|log_2_ FC| > 2, **Supplemental Figures 7a & b**). The comparison between T3 and LM3 presented 83 contrasting proteins. **Supplemental Table 5** lists the top 15 differentially expressed proteins (low & high), including the two strongly expressed oncogenes CD74 and TNC and the clinically applicable biomarker for colorectal liver metastasis CEACAM5.

Furthermore, 19 RNAs in the NM3 vs. T3 and 35 RNAs in the NM3 vs. LM3 comparison were differentially expressed (|log_2_ FC| >1). The comparison between T3 and LM3 observed only nine differentially expressed genes (top genes are presented in **Supplemental Table 6**). Interestingly, only CA1 showed a lower concentration in both malignant tissues compared to adjacent mucosa on RNA and protein levels (RNA level: log_2_ FC = -1.208 and -1.402; protein level: log_2_ FC = -2.170 and -3.133, **Supplemental Figures 7c & d**).

### Patient 4: Female, right-sided CRC (pT4)

Samples of the fourth patient were retrieved during one simultaneous surgery. This opened the possibility of not just collecting one sample of the tumor and its paired liver metastasis but six distinct tumor partitions. Semi-quantitative protein profiling by LC-MS/MS discovered 329 proteins in the NM4 vs. T4 (pooled) and 249 proteins in the NM4 vs. LM4 to be differentially expressed (|log_2_ FC| > 2). Comparing T4 and LM4 presented 155 different expressed proteins (**Supplemental Figures 8a & b**, top proteins are shown in **Supplemental Table 7**). Interestingly, almost all tumor-associated proteins, including the oncogenes DEK and CHD4 and the tumor-suppressor gene MYH9, were lower expressed in LM4 than in T4. By contrast, CEACAM5 was higher expressed in LM4. Particular attention should be drawn to TNC, SRSF3, and OLFM4, which presented the highest protein expression in T4 and LM4 compared to NM4.

After transcriptomic analysis, 35 RNAs indicated a |log_2_ FC| >1 in the NM4 vs. T4 and 61 RNAs in the NM4 vs. LM4 contrast. Comparing T4 vs. LM4 presented only four differentially expressed targets (**Supplemental Figures 8c & d**). **Supplemental Table 8** lists the top differentially expressed RNAs of the three two-group comparisons. The overlap of genes and proteins showed two molecules for the NM4 vs. T4 comparison (OGN, ORM1), one for the NM4 vs. LM4 comparison (CA1), and one for the T4 vs. LM4 comparison (FGG).

### Intra-tumoral proteomic landscape in samples of patient 4

Six distinct parts of the primary tumor of patient 4 were analyzed to assess how different tumor biopsies could display intra-tumoral heterogeneity on the level of the transcriptome and proteome. Interestingly, a Pearson correlation between T4(1-6) vs. NM4 and LM4 vs. NM4 ranged from 0.3596<r<0.7032 on the protein and from 0.2100<r<0.8043 on the transcriptome level (**Supplemental Figures 9 & 10**) highlighting different RNA and protein compositions in the six biopsies. Frequently mutated genes were *ERBB2* (100 %), *APC* (100 %), *GNA11* (100 %), *KRAS* (100 %), *PIK3CA* (100 %), *SMAD4* (100 %), and *TP53* (100 %). All 17 genes with a detectable mutation are presented in **Supplemental Figure 11**.

### Functional enrichment analysis of detected proteins

A gene set enrichment analysis using HALLMARK gene sets was performed to obtain a more comprehensive insight into the biological and functional characteristics of the patient-individual cancer progression. Because of the small effect size, RNA expression data was not used for functional enrichment. With the threshold of FDR < 0.05, the protein expression differences resulted in distinct activated pathways during the normal-to-tumor-to-metastasis transition for the individual patients.

As the pathway analysis of patient 1 revealed xenobiotic metabolism, coagulation, and KRAS signaling as top enriched hallmark gene sets in LM1 compared to NM1 and T1, patient 2 showed activated pathways associated with angiogenesis, coagulation, and the complement system. While proteomic profiling for patient 3 resulted in activated pathways connected to epithelial-mesenchymal transition (EMT), myogenesis, and angiogenesis, patient 4 indicated signatures correlated to oxidative phosphorylation, reactive oxygen species pathway, and bile acid metabolism (**Figure 4**). The most striking result from the data is that (i) single patients employ individual pathways on their route to metastasis, and (ii) the most activated pathways are detected between T and LM comparisons.

**Figure 4:**
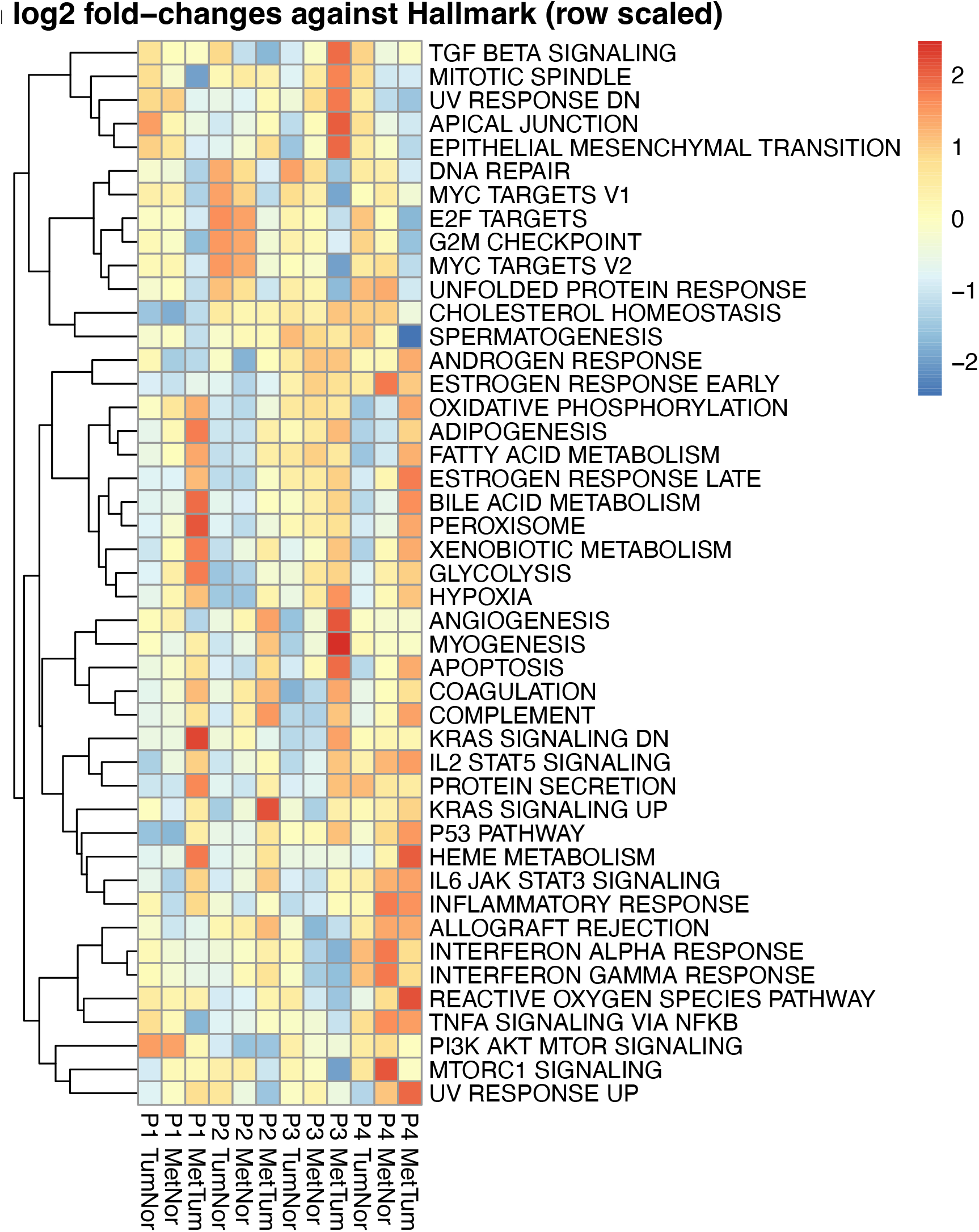
GAGE against HALLMARK gene sets using protein expression data of all two-group comparisons and all patients.

## Discussion

In this report, we describe a panomics investigation of CRCs that integrates a unique set of paired samples of the colon mucosa, primary tumors, and liver metastasis to provide insights into the impact of individual progression. To ensure a robust model and patient comparison, we developed a new protocol to prepare fresh frozen samples with no freeze-and-thaw steps followed by proteomic approaches. This workflow robustly identified potential targets for early detection, diagnosis, or therapeutic intervention.

Our panomics approach indicated differentially expressed genes and proteins comparing adjacent normal mucosa and malignant tissue (T & LM, **Figures 1 & 2**). As one of the few features overlapping between RNA and protein data in the group comparisons, intestinal calcium-activated chloride channel regulator 1 (CLCA1) displayed significantly lower expression in malignant tissue than in healthy colonic mucosa (RNA: log_2_FC_TvsNM_ = -4.02, q-value = 0.0068; log_2_FC_LMvsNM_ = -4.12, q-value = 0.0057; protein: log_2_FC_TvsNM_ = -21.01, q-value = 0.0029; protein log_2_FC_LMvsNM_ = -20.88, q-value = 0.0029). This finding supports evidence from previous observations [33, 34] that shows a decreased expression of CLCA1 during colorectal carcinogenesis. CLCA1 is the gene product of CLCA1 (1p22.3) and is predominantly expressed in intestinal crypts and goblet cells of the colon mucosa. It activates calcium-dependent chloride channels and is primarily involved in mucus regulation [35, 36]. The tumor-suppressive function of CLCA1 is based on the inhibition of EMT and the Wnt/β-catenin signaling pathway: both pathways reduce proliferation but promote differentiation, migration, and invasion [34, 37]. CLCA1 has already been identified as a prognostic factor in colorectal cancer, showing a low expression level associated with poor survival and advanced disease stages [38, 39]. However, the exact mechanisms of decreased expression of CLCA1 in malignant tissues need to be elucidated in more detail. Following the results of CLCA1, the carbonic anhydrase-1 (CA1, 8q21.2) was reported to be decreased in CRC [40] which is consistent with our study (RNA: log_2_FC_TvsNM_ = -4.02, q-value = 0.0068; log_2_FC_LMvsNM_ = -4.12, q-value = 0.0057, protein: log_2_FC_TvsNM_ = -21.01, q-value = 0.0029; log_2_FC_LMvsNM_ = -20.88, q-value = 0.0029). CA1 belongs to the large family of zinc metalloenzymes that maintain pH homeostasis [41] and catalyze the reversible hydration and dehydration reactions of CO_2_/H_2_CO_3_ [42]. Previous studies observed that CA1-mRNA is significantly reduced in colon carcinoma and that a loss of CA1 expression is associated with the disappearance of differentiated epithelial cells [43]. In accordance, mRNA, RNA, and protein levels are reduced in CRC in the data of the human protein atlas [31, 44].

Another important finding was the lower RNA- and protein level detection of matrilin-2 (MATN2) in tumor compared to normal material (RNA: log_2_FC_TvsNM_ = -2.45, q-value = 0.0093; log_2_FCM_LMvsNM_ = -3.71, q-value = 0.0010, protein: log_2_FC_TvsNM_ = -3.75, q-value = 0.0011; log_2_FCM_LMvsNM_ = -4.16, q-value = 0.0004). MATN2 is the largest member of the matrilin family, can bind to fibrillar collagens, and plays a role in cell growth and tissue remodeling [45, 46]. It has been shown that MATN2 expression was significantly altered during cancer progression [47-49].

Most strikingly, we detected patient-individuality and not group affiliation as the primary driver of the transcriptome and proteome in CRC. Each primary carcinoma was closely associated with its metastatic counterpart using cluster algorithms without showing drastic mutational differences (**Figures 1a & 2a**). First, these results are consistent with other studies showing a significant inter-individual heterogeneity of clinicopathologic characteristics in CRC. It has been demonstrated that, e.g., stratification along with right-sided or left-sided tumors, according to the Consensus Molecular Subtypes by Guinney et al., impact the prognosis of the patients [50]. Second, these findings highlight the individual evaluation of patients and the comparison of entity groups. Most biomarkers studies compare two or more groups that compromise specific disease subgroups and/or healthy individuals. However, the expression data observed in this investigation revealed a gain of information by analyzing paired patient samples. While we did not detect any differentially expressed genes and proteins between T- and M-groups, the panomics landscape between individual patient T- and M-samples was drastically changed (**Figures 1 & 2**).

On the specific gene and protein level, we found serine and arginine-rich splicing factor 3 (SRSF3), olfactomedin 4 (OLFM4), and the clinical routine marker CEACAM5 to be differentially expressed during patient-intraindividual colorectal progression. SRSF3 was identified as a differentially expressed protein in patient two and patient 4, showing a high concentration in malignant tissues compared to their adjacent normal mucosa (range log_2_FC_TvsNM_: 2.94 - 6.93; range log_2_FC_LMvsNM_: 4.93 - 4.96). SRSF3 (6p21.31 - p21.2) is a target of the Wnt/β-catenin signaling pathway [51] and exerts its pro-oncogenic effects in different ways [52, 53]. While SRSF3 is co-expressed with the interleukin enhancer-binding factor 3 (ILF3) followed by increased alternative splicing of pro-proliferative ILF3 isoforms [54], it also favors energy production by anaerobic glycolysis due to increased splicing of pyruvate kinase isoform M2 [55].

Next to SRSF3, OLFM4 (13q14.3) showed a prominent expression increase in tumor and metastasis of patient 4 (log_2_ FC_T-NM_: 7.65; log_2_ FC_LM-NM_: 7.65) with no differences between tumor and metastasis (log_2_FC_LMvsT_: 0). OLFM4 is a glycoprotein, involved in cell adhesion processes through cadherin interaction [56], and is predominantly expressed in the crypts of the healthy colonic mucosa [57]. Our finding broadly supports the work of other studies in this area linking OLFM4 over-expression with colorectal disease: van der Flier, Shinozaki, and Huang showed that higher protein levels of OLFM4 are associated with inflammatory bowel diseases, adenomatous precursors, and CRC [57-59].

The detected protein levels of CEACAM5 reflect the challenge of inter- and intra-tumor heterogeneity and the importance of patient individuality. CEACAM5 (synonym for carcinoembryonic antigen, CEA) is over-expressed in about 90% of gastrointestinal, colorectal, and pancreatic cancers [60]. Serum levels of CEA are clinically used to monitor postoperative disease recurrence or response to cancer therapy in colorectal cancer, usually in combination with imaging and endoscopic procedures [61]. The current study found that CEACAM5 is heterogeneously expressed in primary and metastatic colorectal cancer tissue. While a homogenous low protein level of CEACAM5 was detected in normal mucosa, protein expression increases heterogeneously per patient in malignant tissues. In detail, P1 and P4 presented an equal CEA level in both malignant tissues (T & LM), while CEA increased or decreased in LM for patients P2 and P3. This finding is consistent with other studies confirming a strong intra- and inter-individual heterogeneity and limited diagnostic and prognostic usability [62, 63].

In conclusion, we could show that inter- and intra-tumor heterogeneity is detectable by panomics and that rather patients and not disease groups present distinct similarities pointing to individual clinical phenotypes. Next to changes among groups reflecting colorectal cancer progression (e.g., CA1, CLCA1, MATN2), we identified significant expression differences between patient-individual tissues (e.g., SRSF3, OLFM4, CEACAM5) contributing to existing knowledge on patient-specific progression and highlighting the importance of paired samples for precision medicine.

## Data Availability

All data are available online at Gene Expression Omnibus (GEO, https://www.ncbi.nlm.nih.gov/geo/) with the accession number GSE206800 and at ProteomeXchange with the data set identifier PXD036434.

## Abbreviations

2D-DIGE: Two-dimensional gel electrophoresis
CEA/CEACAM5: Carcinoembryonic antigen
CLCA1: calcium-activated chloride channel regulator 1
CRC: Colorectal Cancer
EMT: epithelial-mesenchymal transition
FASP: Filter-aided sample preparation
GEO: Gene Expression Omnibus
H&E: Hematoxylin and eosin
HCD: High-energy collisional dissociation
LC-MS/MS: liquid chromatography-coupled mass spectrometry
LM: Liver metastases
mCRC: metastatic CRC
NGS: Next-generation sequencing
NM: Normal adjacent colon mucosa
OLFM4: Olfactomedin 4
RTA: Real-time analysis
SRSF3: Serine and arginine-rich splicing factor 3
T: Primary colorectal carcinomas
TCGA: The Cancer Genome Atlas
TNC: Tenascin C
tNGS: Targeted amplicon sequencing
TSACP: TruSeq™ Amplicon Cancer Panel

## Acknowledgments

Grants from the Ad Infinitum Foundation and Werner & Clara Kreitz are gratefully acknowledged. This study was performed in collaboration with the Surgical Center for Translational Oncology-Lübeck (SCTO-L), Lübeck Integrated Oncology Network (LION), and the Interdisciplinary Center for Biobanking-Lübeck (ICB-L). The authors would like to thank Katja Klempt-Giessing, Julia Horn, Emma Neumann, and Gisela Grosser-Pape for excellent technical assistance.

