## Supplemental Figures for "Panomics reveals patient-individuality as the major driver for colorectal cancer progression"

**Supplemental Figure 1: Representative images before and after coring**. Section of a tumor sample from P4 in prior to coring, hematoxylin-eosin stained (A). The view direction from R3 is shown schematically. Section of the same sample after coring, hematoxylin-eosin stained (B). The circle with an inner diameter of 1.5 mm marks the punched area. While the dashed line outlines tumorous tissue (T), * marks connective tissue, and → a collection of lymphoid cells.

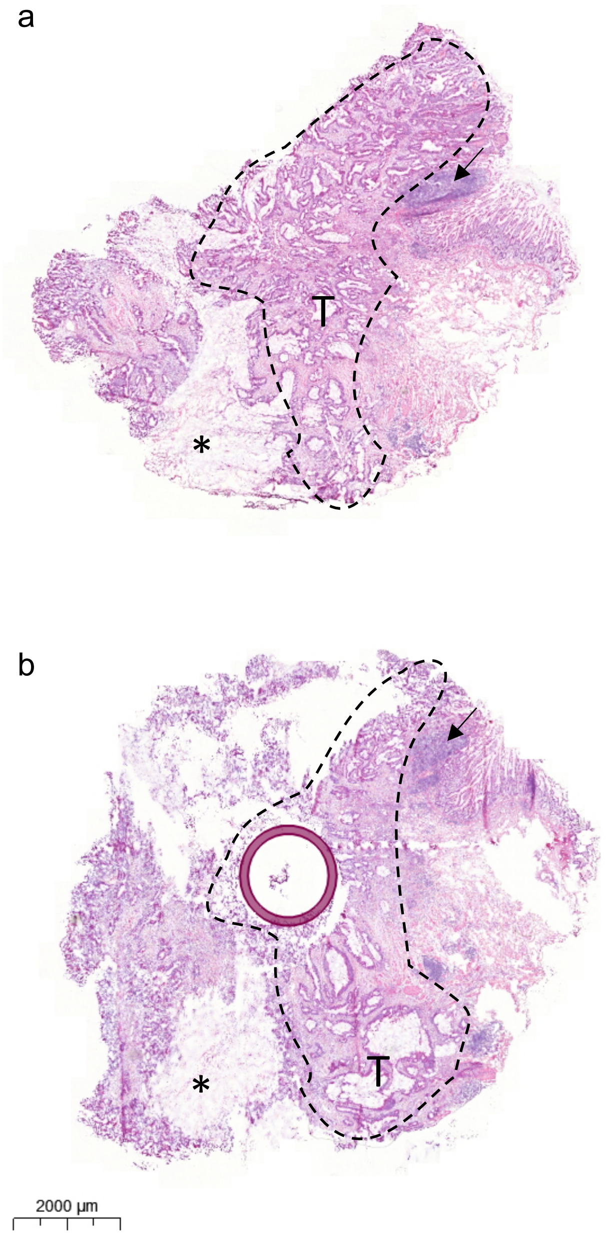

**Supplemental Figure 2: Oncoplot depicting 24 detectable mutated genes in P1-4 sorted and ordered by decreasing frequency.** T, tumor; M, liver metastasis

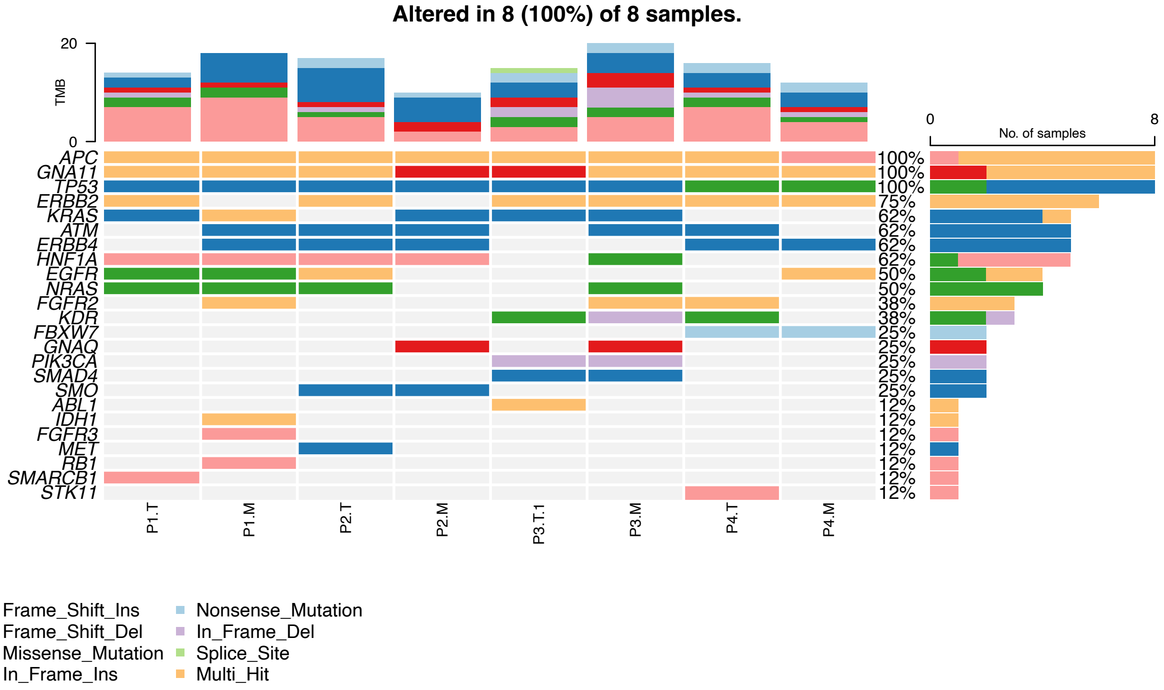

**Supplemental Figure 3: Number of proteins identified and quantified with a 1% FDR in each sample.** Bars indicate the mean and standard deviation. The complete dataset without any missing values contained 2,686 proteins. FDR, False discovery rate, NM, normal mucosa; T, tumor; LM, liver metastasis

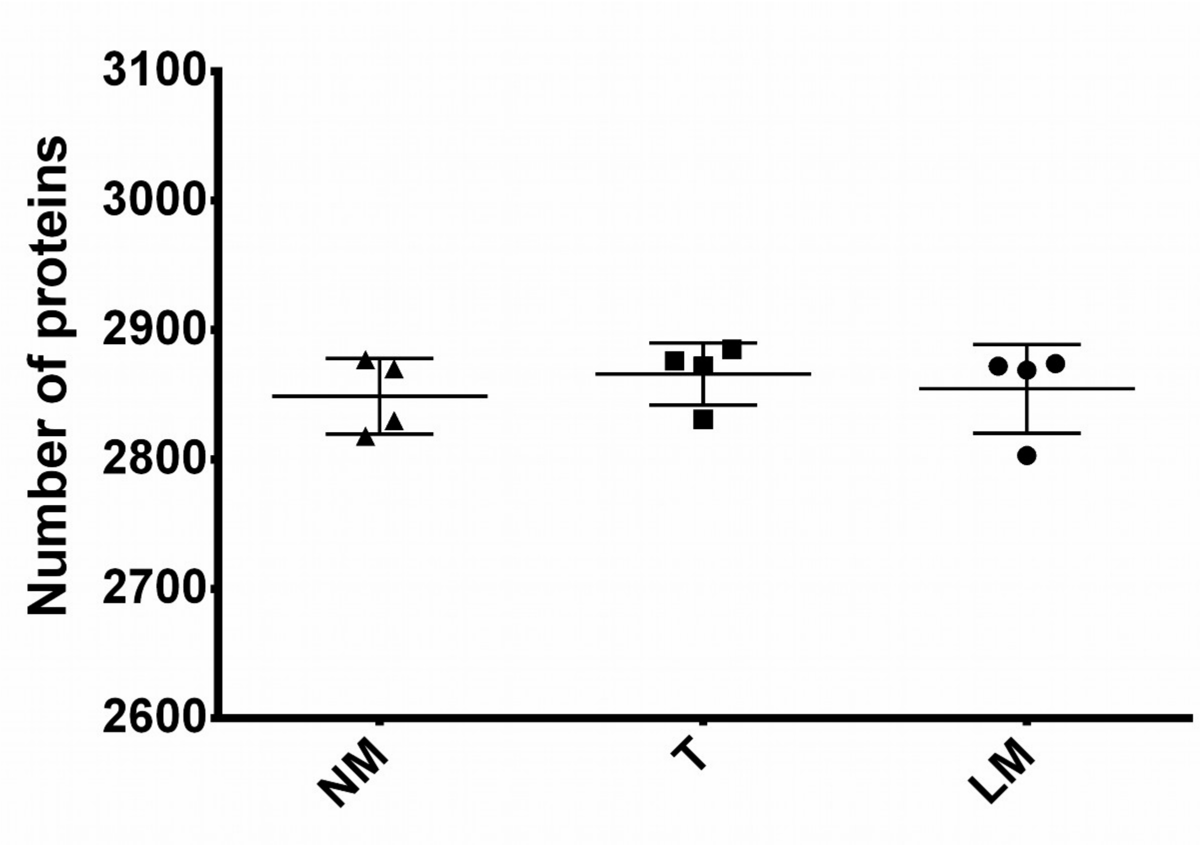

**Supplemental Figure 4: Unsupervised principal component analysis of all three groups after two-dimensional gel electrophoresis.** PCA plot displays all four individual patients (P1, blue; P2, yellow; P3, purple; P4, grey). X- and y-axes show the first and second principal components, respectively.

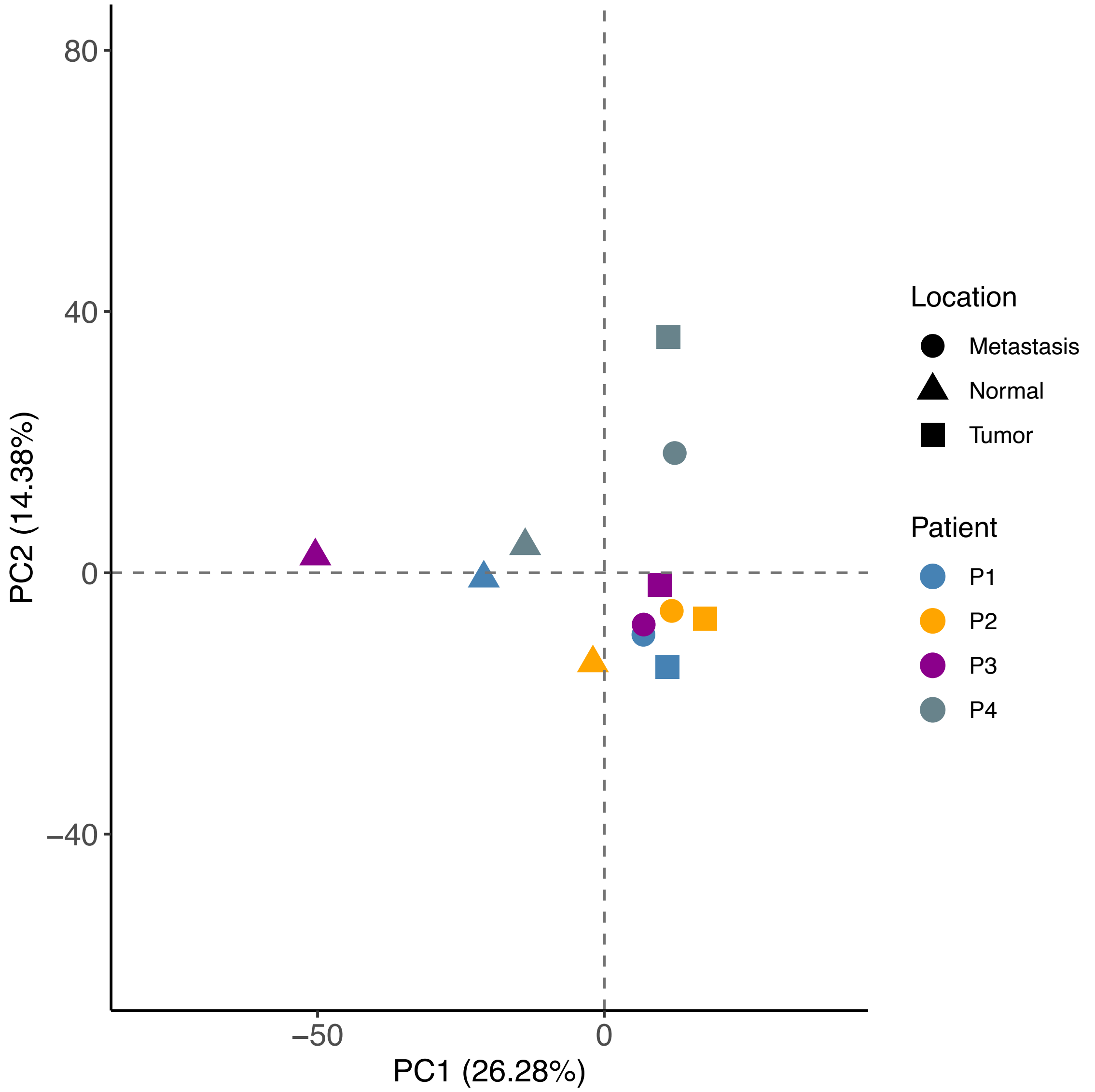

**Supplemental Figure 5: Panomics profiling of patient 1. Fold-change plots showing protein (a, b) and RNA (c, d) expression values for the NM1 vs T1 and NM1 vs LM1 (a, c) as well as for the T1 vs LM1 comparison (b, d).** FC-plots list the top 15 differentially expressed proteins/RNAs with either a higher or lower protein/gene expression, significantly expressed onco-proteins of the group comparisons, and significant targets of the group comparisons overlapping between gene and protein expression data (CD74, TNC, CEACAM5, CA1, CLCA1, MATN2, AHCYL2, FCGBP).

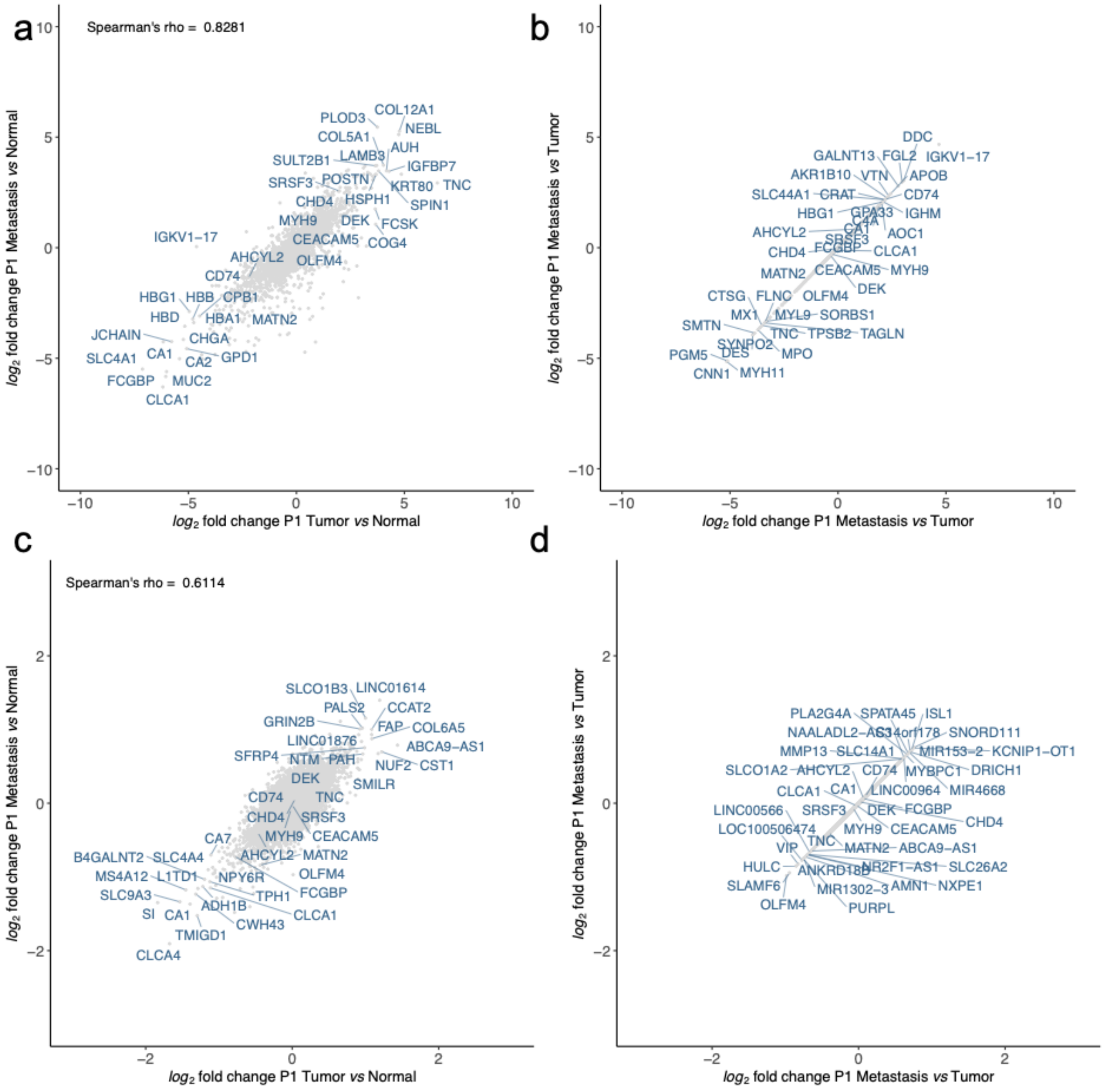

**Supplemental Figure 6: Panomics profiling of patient 2. Fold-change plots showing protein (a, b) and RNA (c, d) expression values for the NM2 vs T2 and NM2 vs LM2 (a, c) as well as for the T2 vs LM2 comparison (b, d).** FC-plots list the top 15 differentially expressed proteins/RNAs with either a higher or lower protein/gene expression, significantly expressed onco-proteins of the group comparisons, and significant targets of the group comparisons overlapping between gene and protein expression data (SRSF3, CEACAM5, CA1, CLCA1, MATN2, AHCYL2, FCGBP).

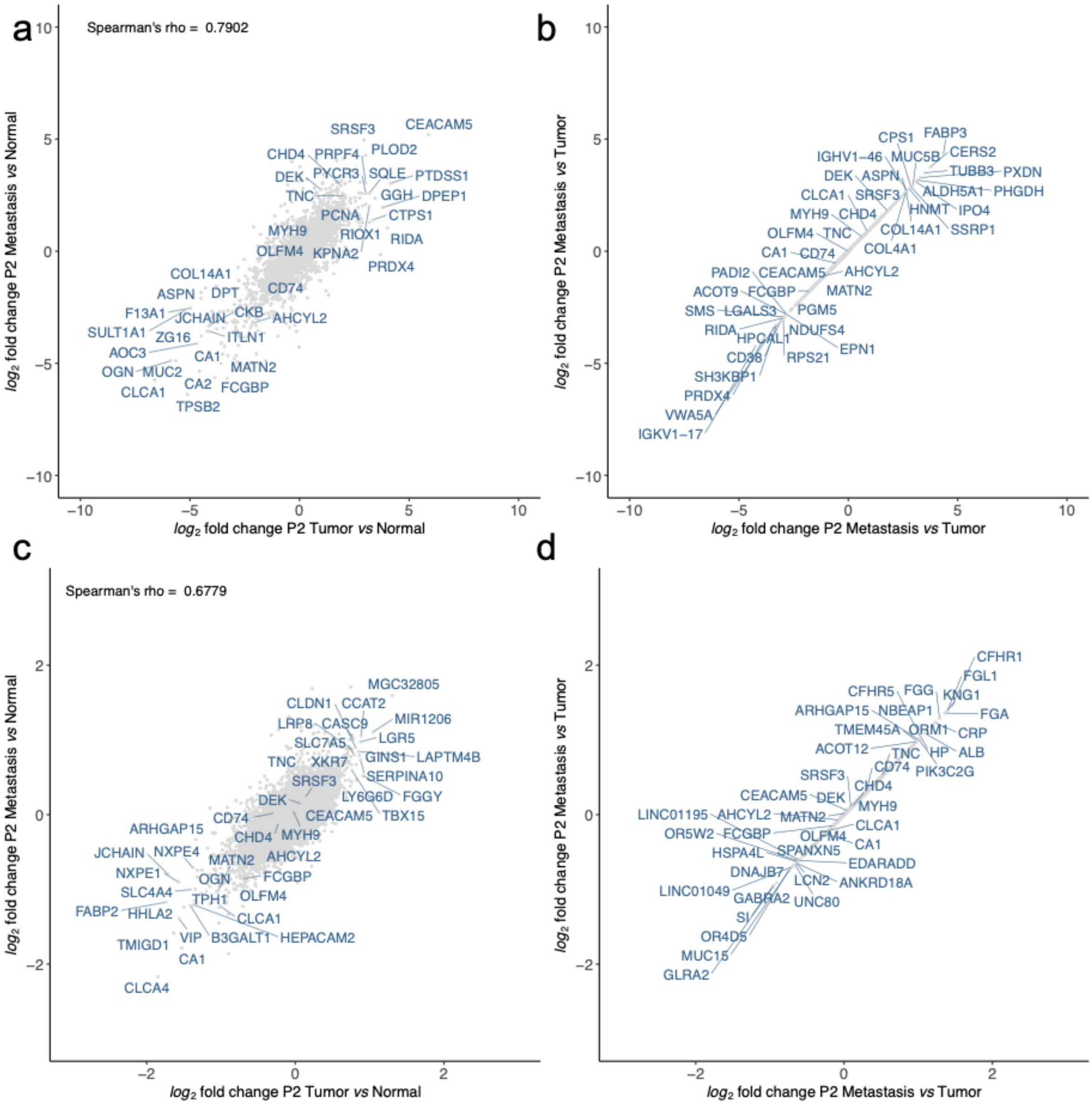

**Supplemental Figure 7: Panomics profiling of patient 3. Fold-change plots showing protein (a, b) and RNA (c, d) expression values for the NM3 vs T3 and NM3 vs LM3 (a, c) as well as for the T3 vs LM3 comparison (b, d).** FC-plots list the top 15 differentially expressed proteins/RNAs with either a higher or lower protein/gene expression, significantly expressed onco-proteins of the group comparisons, and significant targets of the group comparisons overlapping between gene and protein expression data (CD74, TNC, CEACAM5, CA1, CLCA1, MATN2, AHCYL2, FCGBP).

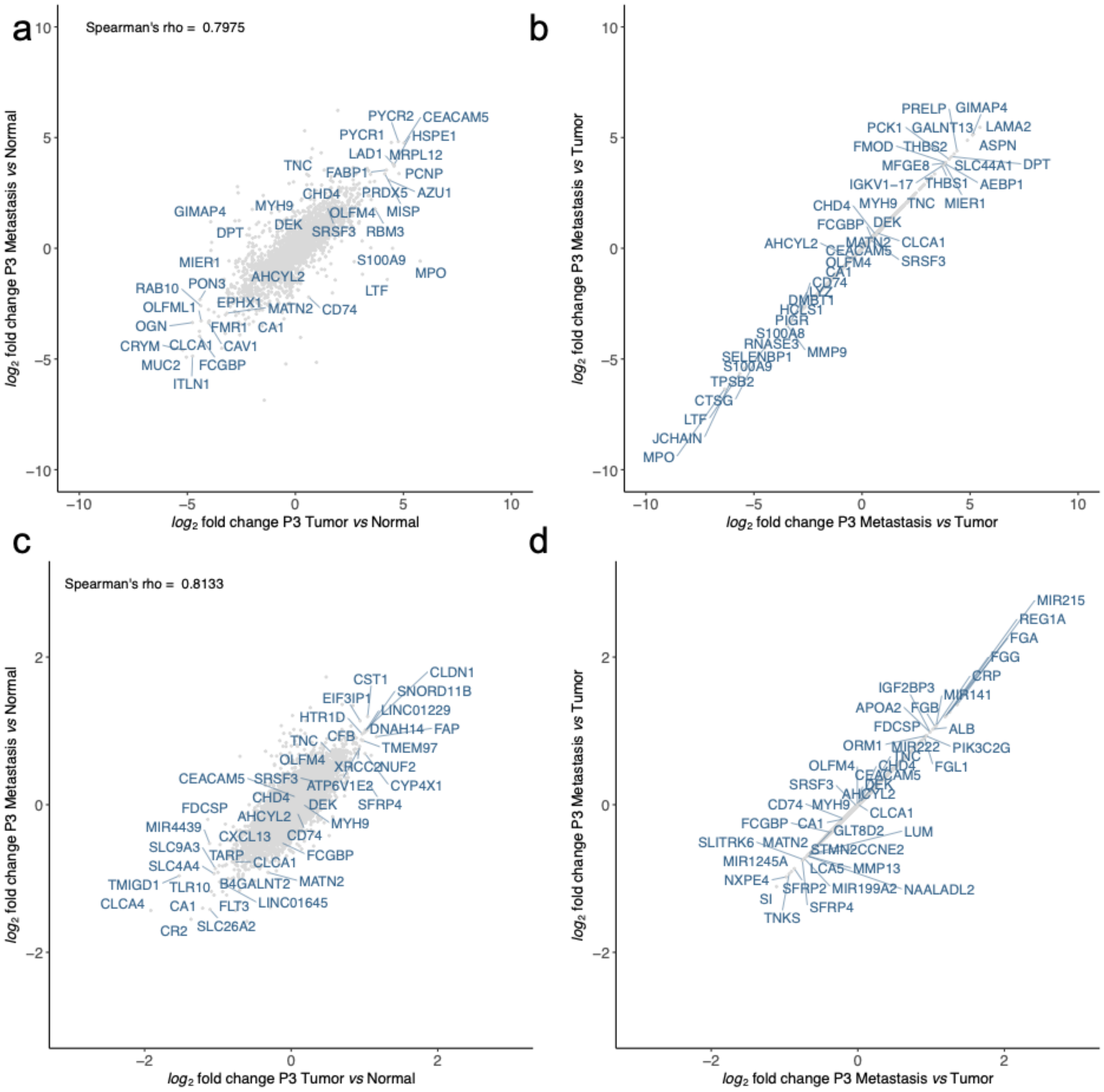

**Supplemental Figure 8: Panomics profiling of patient 4. Fold-change plots showing protein (a, b) and RNA (c, d) expression values for the NM4 vs T4 and NM4 vs LM4 (a, c) as well as for the T4 vs LM4 comparison (b, d).** FC-plots list the top 15 differentially expressed proteins/RNAs with either a higher or lower protein/gene expression, significantly expressed onco-proteins of the group comparisons, and significant targets of the group comparisons overlapping between gene and protein expression data (DEK, CHD4, MYH9, CEACAM5, TNC, SRSF3, OLFM4, CA1, CLCA1, MATN2, AHCYL2, FCGBP).

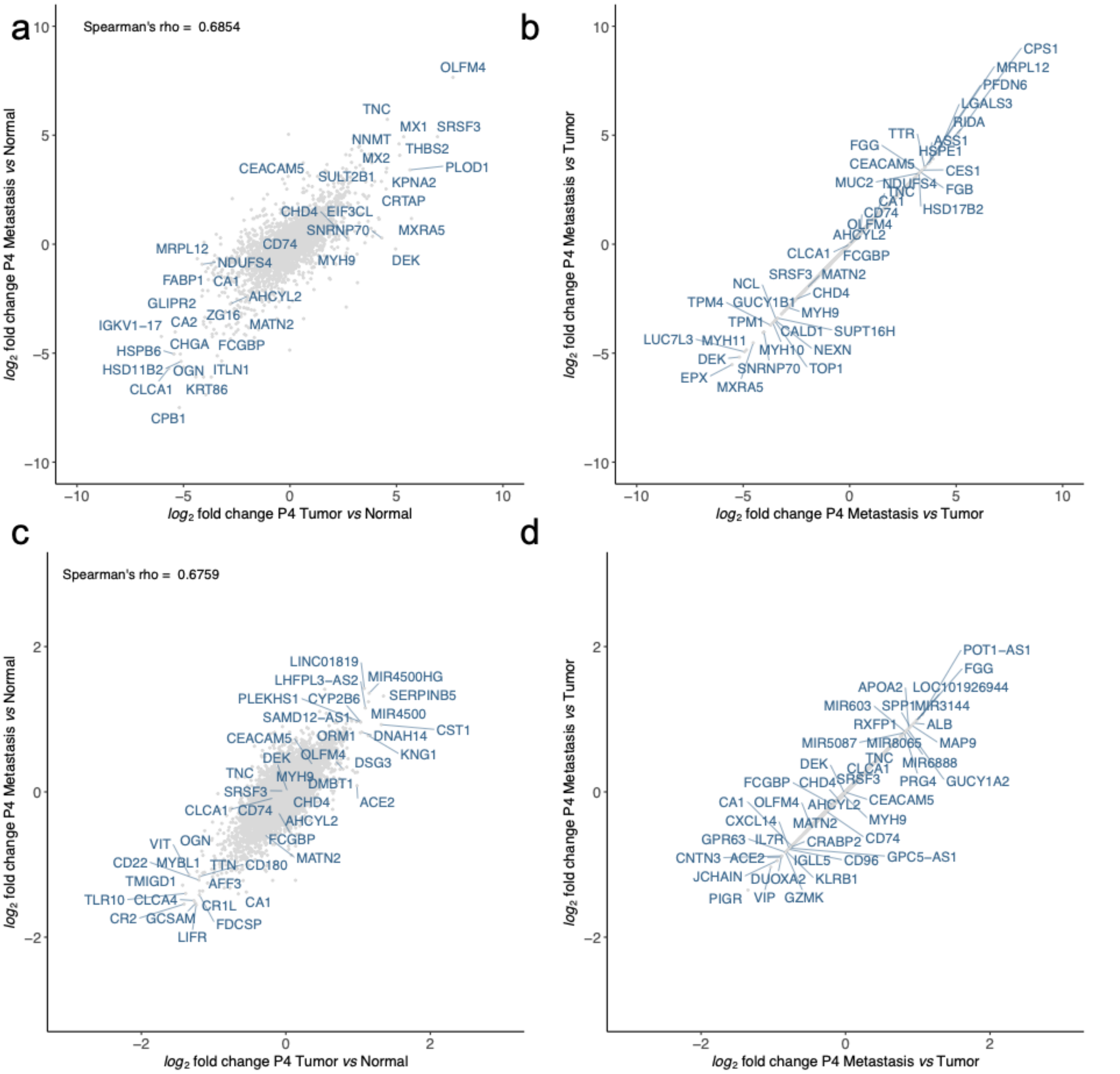

**Supplemental Figure 9: Protein profiling of patient 4 showing six different tumor locations.** Fold-change plots showing protein expression values for the NM vs $T_{1\text{-}6}$ and NM vs LM comparisons. FC-plots list the top 15 differentially expressed proteins with either a higher or lower gene expression, significantly expressed onco-proteins of the group comparisons, and significant targets of the group comparisons overlapping between gene and protein expression data.

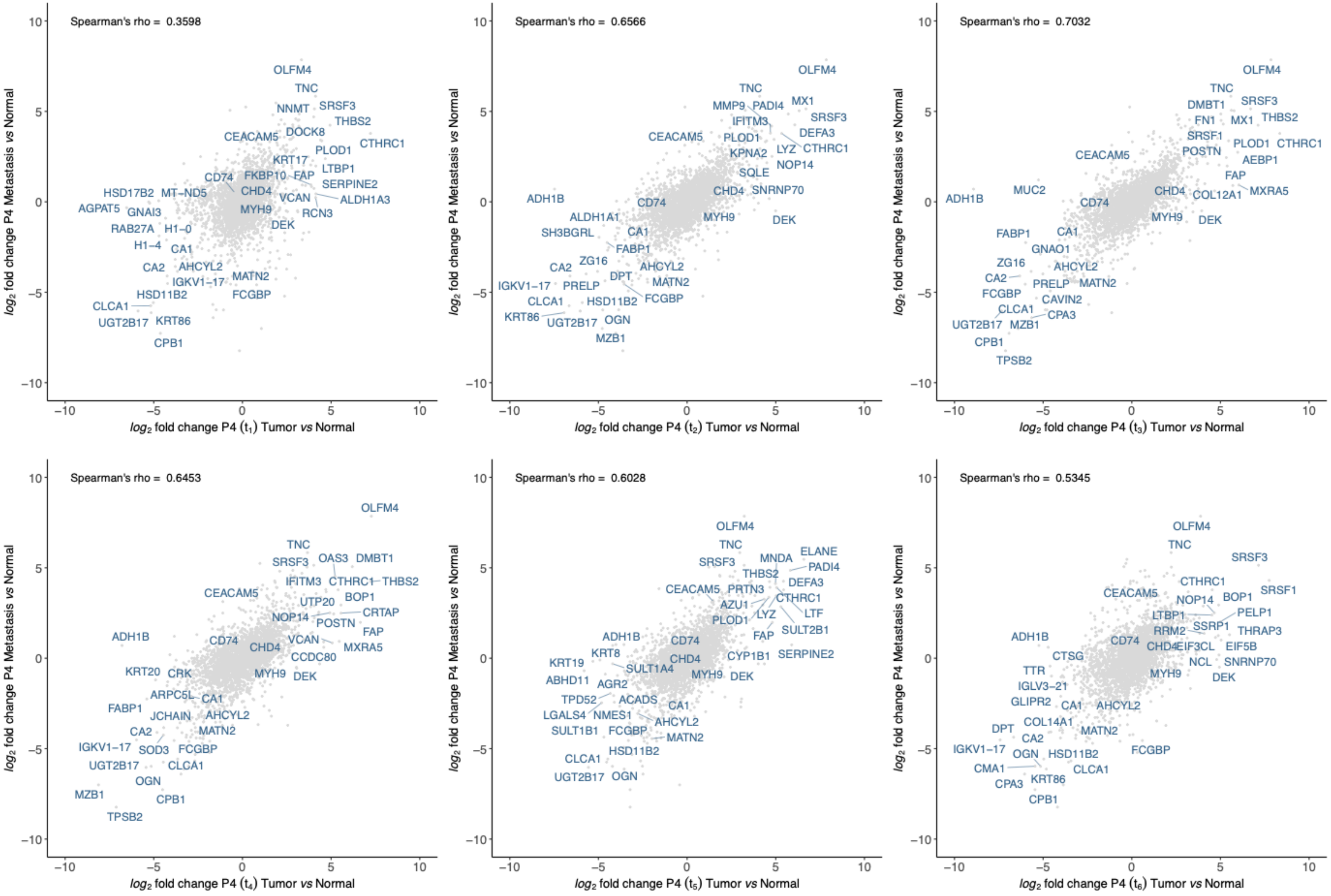

**Supplemental Figure 10: Transcriptomic profiling of patient 4 showing six different tumor locations.** Fold-change plots showing gene expression values for the NM vs $T_{1\text{-}6}$ and NM vs LM comparisons. FC-plots list the top 15 differentially expressed RNAs with either a higher or lower gene expression, significantly expressed onco-proteins of the group comparisons, and significant targets of the group comparisons overlapping between gene and protein expression data.

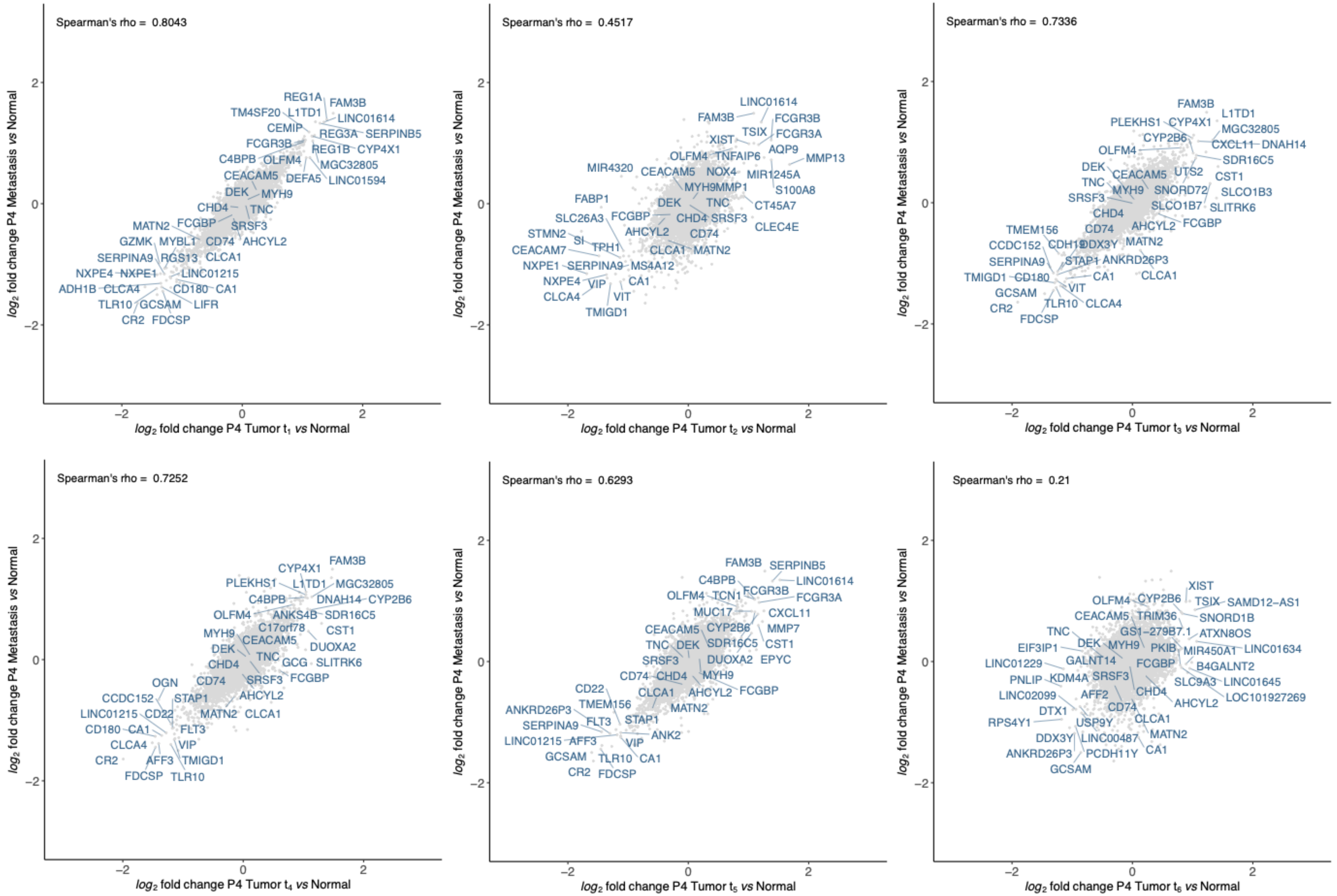

**Supplemental Figure 11*:* Oncoplot depicting 17 detectable mutated genes of different tumor locations in P4 sorted and ordered by decreasing frequency.** T, tumor; M, liver metastasis

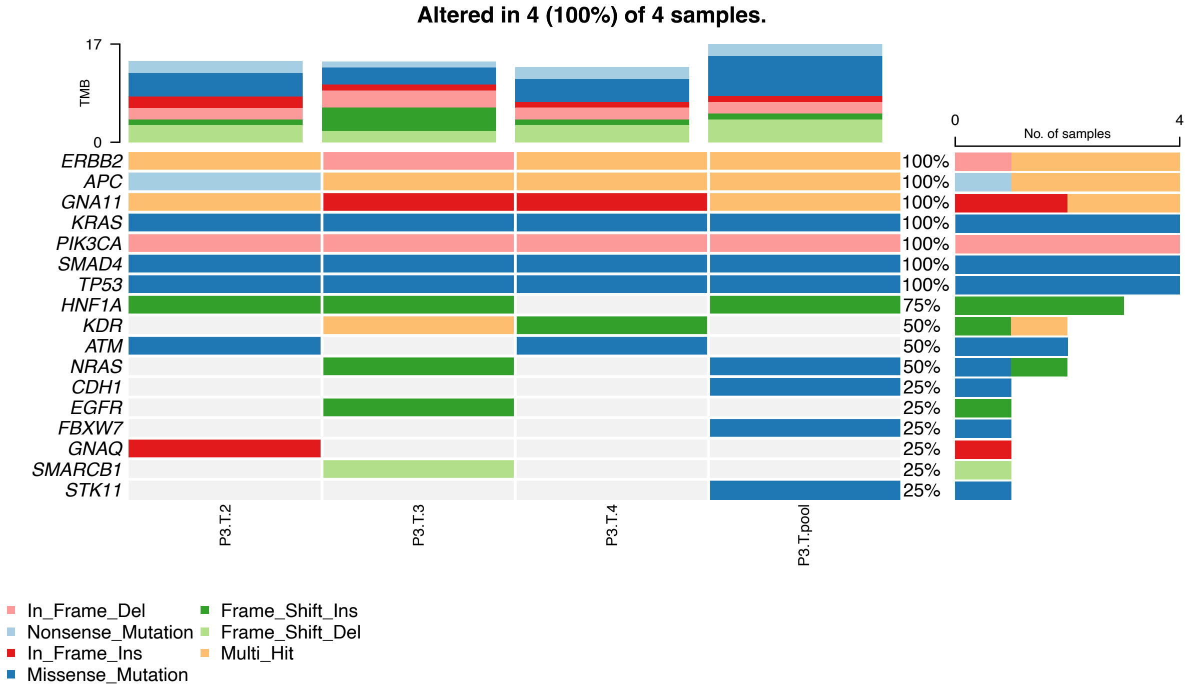
