## Supplemental Tables for "Panomics reveals patient-individuality as the major driver for colorectal cancer progression"

**Supplemental Table 1: Top 30 differentially expressed proteins in two-group comparisons of patient 1.** Listed are the top 15 proteins with increased or decreased expression in the comparison of tumor vs. normal tissue, metastasis vs. normal tissue, and metastasis vs. tumor tissue measured by $log_2$FC. Proteins marked in bold are products of onco- or tumor suppressor genes annotated in NCG 7.0.

| T vs. NM | LM vs. NM | LM vs. T |
|---|---|---|
| **Top 15 proteins with increased expression** | | |
| **TNC** (6.52) | PLOD3 (5.45) | IGKV1-17 (4.67) |
| KRT80 (4.86) | COL12A1 (5.27) | DDC (3.04) |
| COL12A1 (4.77) | NEBL (5.12) | FGL2 (2.97) |
| NEBL (4.74) | AEBP1 (4.29) | GALNT13 (2.79) |
| IGFBP7 (4.28) | **SYK** (4.17) | APOB (2.44) |
| AUH (4.17) | NQO1 (4.02) | VTN (2.35) |
| COL5A1 (4.03) | S100A11 (3.98) | AKR1B10 (2.26) |
| SPIN1 (3.81) | THBS1 (3.97) | **CD74** (2.18) |
| PLOD3 (3.75) | COL5A1 (3.73) | CRAT (2.17) |
| LAMB3 (3.72) | LAMB3 (3.72) | SLC44A1 (2.17) |
| HSPH1 (3.69) | SULT2B1 (3.71) | IGHM (2.08) |
| COG4 (3.67) | TGFBI (3.70) | HBG1 (2.07) |
| FCSK (3.65) | PDP1 (3.61) | AOC1 (2.03) |
| POSTN (3.65) | MEP1A (3.52) | GPA33 (2.00) |
| SULT2B1 (3.65) | AUH (3.48) | C4A (1.91) |
| **Top 15 proteins with decreased expression** | | |
| SLC4A1 (-7.13) | CLCA1 (-6.30) | MYH11 (-5.15) |
| CLCA1 (-6.18) | FCGBP (-5.82) | CNN1 (-5.13) |
| CA1 (-6.17) | MUC2 (-5.60) | PGM5 (-5.12) |
| FCGBP (-6.05) | SLC4A1 (-5.49) | DES (-3.95) |
| MUC2 (-6.01) | FABP4 (-5.05) | SYNPO2 (-3.87) |
| JCHAIN (-5.78) | CA2 (-5.02) | SMTN (-3.86) |
| CA2 (-5.41) | PIGR (-4.84) | MPO (-3.71) |
| CHGA (-5.23) | CAV1 (-4.83) | MX1 (-3.63) |
| HBD (-5.17) | PGM5 (-4.73) | **TNC** (-3.59) |
| GPD1 (-5.08) | FHL1 (-4.70) | CTSG (-3.52) |
| HBG1 (-4.96) | GPD1 (-4.57) | TPSB2 (-3.51) |
| HBB (-4.80) | CAVIN2 (-4.41) | TAGLN (-3.40) |
| HBA1 (-4.75) | EPX (-4.38) | FLNC (-3.37) |
| IGKV1-17 (-4.63) | MYH11 (-4.27) | SORBS1 (-3.33) |
| CPB1 (-4.48) | CA1 (-4.26) | MYL9 (-3.30) |

FC, fold change; NCG, Network of Cancer Genes, NM, normal mucosa; T, tumor; LM, liver metastasis

**Supplemental Table 2: Top 30 differentially expressed RNAs in two-group comparisons of patient 1.** Listed are the top 15 RNAs with increased or decreased expression in the comparison of tumor vs. normal tissue, metastasis vs. normal tissue, and metastasis vs. tumor tissue measured by $log_2FC$. RNAs marked in bold are onco- or tumor suppressor genes annotated in NCG 7.0.

| T vs. NM | LM vs. NM | LM vs. T |
|---|---|---|
| **Top 15 RNAs with increased expression** | | |
| ABCA9-AS1 (1.44) | LINC01614 (1.40) | SNORD111 (0.76) |
| CST1 (1.22) | SLCO1B3 (1.16) | KCNIP1-OT1 (0.75) |
| LINC01614 (1.19) | IL6 (1.11) | ISL1 (0.72) |
| NUF2 (1.17) | PALS2 (1.02) | MIR153-2 (0.72) |
| COL6A5 (1.09) | GRIN2B (1.01) | C14orf178 (0.70) |
| CCAT2 (1.09) | CCAT2 (1.00) | DRICH1 (0.70) |
| FAP (1.08) | MIR6888 (0.93) | MIR4668 (0.69) |
| SLCO1B3 (1.00) | FAP (0.93) | SPATA45 (0.68) |
| **SFRP4** (0.99) | MIR548AR (0.93) | PLA2G4A (0.66) |
| NTM (0.96) | SLC14A1 (0.92) | MYBPC1 (0.65) |
| PALS2 (0.96) | LOC101927967 (0.90) | NAALADL2-AS3 (0.62) |
| LINC01876 (0.95) | CLDN10-AS1 (0.88) | LINC00964 (0.62) |
| PAH (0.94) | COL6A5 (0.88) | SLC14A1 (0.61) |
| GRIN2B (0.92) | SERPINE1 (0.84) | MMP13 (0.60) |
| SMILR (0.92) | LINC01876 (0.84) | SLCO1A2 (0.60) |
| **Top 15 RNAs with decreased expression** | | |
| SI (-1.84) | CLCA4 (-1.91) | OLFM4 (-0.98) |
| CLCA4 (-1.68) | TMIGD1 (-1.52) | SLAMF6 (-0.94) |
| SLC9A3 (-1.53) | SLC26A2 (-1.48) | HULC (-0.85) |
| MS4A12 (-1.45) | VIP (-1.40) | VIP (-0.83) |
| CA1 (-1.39) | CA1 (-1.37) | LOC100506474 (-0.77) |
| CWH43 (-1.32) | SI (-1.35) | PURPL (-0.75) |
| TMIGD1 (-1.30) | SLC9A3 (-1.34) | MIR1302-3 (-0.75) |
| L1TD1 (-1.29) | HEPACAM2 (-1.29) | ANKRD18B (-0.74) |
| ADH1B (-1.22) | PIGR (-1.28) | AMN1 (-0.73) |
| B4GALNT2 (-1.20) | CWH43 (-1.23) | NXPE1 (-0.70) |
| SLC4A4 (-1.15) | NXPE1 (-1.23) | NR2F1-AS1 (-0.70) |
| TPH1 (-1.12) | MS4A12 (-1.18) | SLC26A2 (-0.69) |
| CLCA1 (-1.12) | REP15 (-1.16) | LINC00566 (-0.67) |
| CA7 (-1.11) | L1TD1 (-1.16) | ABCA9-AS1 (-0.65) |
| NPY6R (-1.09) | CLCA1 (-1.15) | **TNC** (-0.64) |

FC, fold change; NCG, Network of Cancer Genes, NM, normal mucosa; T, tumor; LM, liver metastasis

**Supplemental Table 3: Top 30 differentially expressed proteins in two-group comparisons of patient 2.** Listed are the top 15 proteins with increased or decreased expression in the comparison of tumor vs. normal tissue, metastasis vs. normal tissue, and metastasis vs. tumor tissue measured by $log_2FC$. Proteins marked in bold are products of onco- or tumor suppressor genes annotated in NCG 7.0.

| T vs. NM | LM vs. NM | LM vs. T |
|---|---|---|
| **Top 15 proteins with increased expression** | | |
| CEACAM5 (5.89) | IPO4 (5.53) | FABP3 (4.35) |
| PTDSS1 (4.16) | CEACAM5 (5.19) | CERS2 (3.75) |
| RIDA (4.01) | **SRSF3** (4.96) | TUBB3 (3.50) |
| DPEP1 (3.79) | PLOD2 (4.28) | PXDN (3.28) |
| PRDX4 (3.69) | SUPT16H (4.15) | PHGDH (3.17) |
| GGH (3.45) | PXDN (4.05) | ALDH5A1 (3.12) |
| CTPS1 (3.34) | **CNBP** (4.04) | IPO4 (3.09) |
| RIOX1 (3.20) | FABP3 (3.99) | MUC5B (2.96) |
| SQLE (3.19) | CERS2 (3.77) | SSRP1 (2.87) |
| KPNA2 (3.19) | UTP20 (3.61) | CPS1 (2.87) |
| PYCR3 (3.03) | HDAC2 (3.43) | HNMT (2.79) |
| PCNA (3.02) | AGXT (3.35) | IGHV1-46 (2.69) |
| PLOD2 (3.02) | BYSL (3.31) | ASPN (2.65) |
| PRPF4 (3.01) | AEBP1 (3.31) | COL14A1 (2.65) |
| **SRSF3** (2.94) | DDX27 (3.27) | COL4A1 (2.64) |
| **Top 15 proteins with decreased expression** | | |
| CLCA1 (-6.61) | TPSB2 (-6.39) | IGKV1-17 (-4.25) |
| OGN (-5.89) | CLCA1 (-5.74) | PRDX4 (-3.84) |
| MUC2 (-5.65) | CHGA (-5.68) | HPCAL1 (-3.44) |
| SULT1A1 (-5.27) | FCGBP (-5.64) | VWA5A (-3.33) |
| TPSB2 (-5.11) | CA2 (-5.35) | SH3KBP1 (-3.27) |
| F13A1 (-4.97) | CPA3 (-5.32) | CD38 (-3.09) |
| AOC3 (-4.69) | PGM5 (-5.05) | RPS21 (-2.98) |
| CA2 (-4.57) | CRYAB (-5.01) | RIDA (-2.97) |
| ASPN (-4.49) | PADI2 (-4.99) | NDUFS4 (-2.94) |
| DPT (-4.49) | KRT86 (-4.94) | SMS (-2.93) |
| ZG16 (-4.47) | OGN (-4.89) | LGALS3 (-2.84) |
| COL14A1 (-4.29) | VWA5A (-4.89) | ACOT9 (-2.82) |
| JCHAIN (-4.21) | MUC2 (-4.89) | PADI2 (-2.82) |
| CKB (-4.14) | SULT1B1 (-4.80) | EPN1 (-2.81) |
| ITLN1 (-4.11) | CAVIN2 (-4.66) | PGM5 (-2.64) |

FC, fold change; NCG, Network of Cancer Genes, NM, normal mucosa; T, tumor; LM, liver metastasis

**Supplemental Table 4: Top 30 differentially expressed RNAs in two-group comparisons of patient 2.** Listed are the top 15 RNAs with increased or decreased expression in the comparison of tumor vs. normal tissue, metastasis vs. normal tissue, and metastasis vs. tumor tissue measured by $log_2FC$. RNAs marked in bold are onco- or tumor suppressor genes annotated in NCG 7.0.

| T vs. NM | LM vs. NM | LM vs. T |
|---|---|---|
| **Top 15 RNAs with increased expression** | | |
| MGC32805 (1.30) | APOA2 (1.71) | CFHR1 (1.46) |
| MIR1206 (1.04) | CFHR1 (1.69) | FGL1 (1.40) |
| FGGY (0.91) | MGC32805 (1.59) | KNG1 (1.39) |
| CCAT2 (0.89) | ORM1 (1.47) | FGA (1.36) |
| SERPINA10 (0.88) | FGL1 (1.31) | FGG (1.29) |
| LGR5 (0.88) | CRP (1.22) | NBEAP1 (1.24) |
| GINS1 (0.86) | FGG (1.22) | CRP (1.23) |
| LAPTM4B (0.85) | FGB (1.20) | ORM1 (1.20) |
| LY6G6D (0.82) | ALB (1.17) | ALB (1.09) |
| XKR7 (0.82) | CFHR2 (1.17) | HP (1.04) |
| CLDN1 (0.81) | FGA (1.14) | CFHR5 (1.03) |
| SLC7A5 (0.79) | HP (1.12) | PIK3C2G (1.01) |
| CASC9 (0.78) | MIR1206 (1.11) | ARHGAP15 (1.01) |
| TBX15 (0.76) | ABCA9-AS1 (1.10) | TMEM45A (0.98) |
| LRP8 (0.76) | ORM2 (1.09) | ACOT12 (0.98) |
| **Top 15 RNAs with decreased expression** | | |
| CLCA4 (-1.85) | CLCA4 (-2.17) | SI (-0.96) |
| FABP2 (-1.74) | SI (-1.86) | GLRA2 (-0.94) |
| JCHAIN (-1.72) | CA1 (-1.78) | GABRA2 (-0.91) |
| TMIGD1 (-1.64) | TMIGD1 (-1.58) | OR4D5 (-0.75) |
| NXPE1 (-1.57) | SULT1B1 (-1.45) | MUC15 (-0.72) |
| VIP (-1.56) | VIP (-1.39) | LINC01049 (-0.70) |
| CA1 (-1.53) | SLITRK6 (-1.33) | DNAJB7 (-0.69) |
| HHLA2 (-1.50) | ABCG2 (-1.24) | UNC80 (-0.66) |
| B3GALT1 (-1.41) | HHLA2 (-1.24) | LCN2 (-0.65) |
| SLC4A4 (-1.41) | PKIB (-1.23) | ANKRD18A (-0.65) |
| NXPE4 (-1.38) | CLCA1 (-1.23) | HSPA4L (-0.63) |
| HEPACAM2 (-1.37) | B3GALT1 (-1.22) | OR5W2 (-0.62) |
| TPH1 (-1.36) | HEPACAM2 (-1.21) | EDARADD (-0.61) |
| OGN (-1.35) | MUC2 (-1.19) | LINC01195 (-0.61) |
| ARHGAP15 (-1.34) | MIR8065 (-1.18) | SPANXN5 (-0.60) |

FC, fold change; NCG, Network of Cancer Genes, NM, normal mucosa; T, tumor; LM, liver metastasis

**Supplemental Table 5: Top 30 differentially expressed proteins in two-group comparisons of patient 3.** Listed are the top 15 proteins with increased or decreased expression in the comparison of tumor vs. normal tissue, metastasis vs. normal tissue, and metastasis vs. tumor tissue measured by $log_2FC$. Proteins marked in bold are products of onco- or tumor suppressor genes annotated in NCG 7.0.

| T vs. NM | LM vs. NM | LM vs. T |
|---|---|---|
| **Top 15 proteins with increased expression** | | |
| MPO (5.78) | THBS2 (6.23) | LAMA2 (5.46) |
| CEACAM5 (5.03) | GALNT13 (5.31) | ASPN (5.16) |
| PCNP (4.80) | PYCR2 (4.82) | GIMAP4 (5.12) |
| PYCR2 (4.76) | PYCR1 (4.78) | GALNT13 (4.88) |
| HSPE1 (4.57) | CEACAM5 (4.75) | PRELP (4.40) |
| MRPL12 (4.57) | THBS1 (4.49) | THBS2 (4.26) |
| LAD1 (4.51) | COL12A1 (4.31) | DPT (4.14) |
| S100A9 (4.50) | GPX2 (4.24) | SLC44A1 (4.04) |
| PYCR1 (4.44) | IGFBP7 (4.08) | PCK1 (4.03) |
| AZU1 (4.35) | AEBP1 (3.92) | FMOD (3.87) |
| LTF (4.25) | LAD1 (3.82) | MFGE8 (3.85) |
| FABP1 (4.21) | HSPE1 (3.77) | AEBP1 (3.83) |
| MISP (4.14) | MRPL12 (3.71) | MIER1 (3.82) |
| RBM3 (3.69) | BAIAP2L1 (3.64) | IGKV1-17 (3.75) |
| PRDX5 (3.66) | SMAP (3.60) | THBS1 (3.71) |
| **Top 15 proteins with decreased expression** | | |
| CRYM (-5.21) | TPSB2 (-6.86) | MPO (-6.36) |
| MUC2 (-5.05) | CTSG (-5.22) | JCHAIN (-6.13) |
| OGN (-4.76) | JCHAIN (-5.15) | LTF (-5.66) |
| ITLN1 (-4.76) | CPA3 (-5.07) | TPSB2 (-5.42) |
| CLCA1 (-4.44) | MUC2 (-4.92) | CTSG (-4.79) |
| FCGBP (-4.43) | ITLN1 (-4.87) | S100A9 (-4.50) |
| PON3 (-4.42) | CRYM (-4.60) | SELENBP1 (-4.28) |
| RAB10 (-4.38) | PGM5 (-4.52) | RNASE3 (-4.26) |
| OLFML1 (-4.36) | ADAM10 (-4.27) | S100A8 (-3.73) |
| CAV1 (-4.03) | RNASE3 (-4.03) | MMP9 (-3.47) |
| EPHX1 (-4.02) | FCGBP (-3.98) | PIGR (-3.33) |
| FMR1 (-3.99) | SLC4A1 (-3.91) | LYZ (-3.28) |
| GIMAP4 (-3.92) | F13A1 (-3.85) | HCLS1 (-3.28) |
| MIER1 (-3.91) | CLCA1 (-3.75) | **CD74** (-2.82) |
| DPT (-3.87) | RAB1A (-3.52) | DMBT1 (-2.76) |

FC, fold change; NCG, Network of Cancer Genes, NM, normal mucosa; T, tumor; LM, liver metastasis

**Supplemental Table 6: Top 30 differentially expressed RNAs in two-group comparisons of patient 3.** Listed are the top 15 RNAs with increased or decreased expression in the comparison of tumor vs. normal tissue, metastasis vs. normal tissue, and metastasis vs. tumor tissue measured by $log_2FC$. RNAs marked in bold are onco- or tumor suppressor genes annotated in NCG 7.0.

| T vs. NM | LM vs. NM | LM vs. T |
|---|---|---|
| **Top 15 RNAs with increased expression** | | |
| FAP (1.16) | REG1A (1.73) | MIR215 (1.35) |
| NUF2 (1.09) | SERPINB5 (1.36) | REG1A (1.25) |
| **SFRP4** (1.05) | NMUR2 (1.34) | FGA (1.21) |
| CST1 (1.04) | FGB (1.31) | FGG (1.19) |
| SNORD11B (1.04) | FGG (1.21) | CRP (1.07) |
| CLDN1 (1.01) | CST1 (1.20) | MIR141 (1.07) |
| CYP4X1 (1.01) | LINC01610 (1.17) | FGB (1.05) |
| LINC01229 (1.01) | ORM1 (1.14) | ALB (1.02) |
| DNAH14 (1.00) | EIF3IP1 (1.14) | IGF2BP3 (0.99) |
| TMEM97 (0.98) | FGA (1.07) | APOA2 (0.98) |
| HTR1D (0.97) | VNN1 (1.07) | FDCSP (0.94) |
| CFB (0.96) | MIR5087 (1.02) | ORM1 (0.94) |
| EIF3IP1 (0.94) | APOA2 (1.02) | PIK3C2G (0.93) |
| XRCC2 (0.93) | CLDN1 (1.01) | FGL1 (0.91) |
| ATP6V1E2 (0.92) | MIR301A (1.01) | MIR222 (0.90) |
| **Top 15 RNAs with decreased expression** | | |
| CLCA4 (-1.92) | NXPE4 (-1.58) | SI (-1.11) |
| TMIGD1 (-1.53) | CR2 (-1.55) | NXPE4 (-0.98) |
| CR2 (-1.37) | STMN2 (-1.44) | TNKS (-0.94) |
| CA1 (-1.21) | CLCA4 (-1.43) | MIR1245A (-0.91) |
| FDCSP (-1.14) | SLC26A2 (-1.42) | SFRP2 (-0.87) |
| MIR4439 (-1.11) | CA1 (-1.40) | **SFRP4** (-0.76) |
| SLC26A2 (-1.11) | SI (-1.39) | SLITRK6 (-0.74) |
| TLR10 (-1.09) | TRHDE (-1.33) | MIR199A2 (-0.73) |
| **FLT3** (-1.05) | **FLT3** (-1.22) | LCA5 (-0.70) |
| SLC4A4 (-1.05) | MS4A12 (-1.22) | NAALADL2 (-0.69) |
| SLC9A3 (-1.04) | VIP (-1.18) | MMP13 (-0.69) |
| B4GALNT2 (-1.00) | TLR10 (-1.18) | CCNE2 (-0.68) |
| TARP (-0.98) | TMEM236 (-1.16) | LUM (-0.68) |
| LINC01645 (-0.98) | SNORD114-12 (-1.05) | STMN2 (-0.68) |
| CXCL13 (-0.97) | LINC01645 (-1.04) | GLT8D2 (-0.67) |

FC, fold change; NCG, Network of Cancer Genes, NM, normal mucosa; T, tumor; LM, liver metastasis

**Supplemental Table 7: Top 30 differentially expressed proteins in two-group comparisons of patient 4.** Listed are the top 15 proteins with increased or decreased expression in the comparison of tumor vs. normal tissue, metastasis vs. normal tissue, and metastasis vs. tumor tissue measured by $log_2$FC. Proteins marked in bold are products of onco- or tumor suppressor genes annotated in NCG 7.0.

| T vs. NM | LM vs. NM | LM vs. T |
|---|---|---|
| **Top 15 proteins with increased expression** | | |
| OLFM4 (7.65) | OLFM4 (7.65) | CPS1 (5.11) |
| **SRSF3** (6.93) | **TNC** (5.73) | RIDA (4.80) |
| MXRA5 (5.70) | CPS1 (5.04) | LGALS3 (4.12) |
| PLOD1 (5.63) | **SRSF3** (4.93) | HSPE1 (3.91) |
| MX1 (5.34) | MX1 (4.93) | ASS1 (3.78) |
| THBS2 (5.15) | DMBT1 (4.83) | MRPL12 (3.69) |
| NNMT (5.12) | NNMT (4.60) | PFDN6 (3.55) |
| **DEK** (4.96) | ISG15 (4.51) | TTR (3.52) |
| **TNC** (4.57) | FN1 (4.46) | CES1 (3.40) |
| MX2 (4.54) | OAS3 (4.35) | FGG (3.35) |
| KPNA2 (4.51) | MUC1 (4.29) | CEACAM5 (3.34) |
| CRTAP (4.51) | MMP9 (4.13) | FGB (3.31) |
| SNRNP70 (4.31) | HK3 (4.11) | MUC2 (3.26) |
| SULT2B1 (4.29) | THBS1 (4.08) | HSD17B2 (3.26) |
| EIF3CL (4.18) | THBS2 (4.08) | NDUFS4 (3.21) |
| **Top 15 proteins with decreased expression** | | |
| IGKV1-17 (-6.04) | CPB1 (-7.50) | EPX (-5.52) |
| CLCA1 (-5.69) | TPSB2 (-6.91) | **DEK** (-5.18) |
| HSPB6 (-5.45) | ITLN1 (-6.10) | LUC7L3 (-4.94) |
| CA2 (-5.39) | CPA1 (-6.10) | MYH11 (-4.85) |
| CPB1 (-5.20) | KRT86 (-6.09) | MXRA5 (-4.53) |
| CHGA (-5.15) | CLCA1 (-5.70) | TPM1 (-4.06) |
| HSD11B2 (-5.12) | OGN (-5.65) | SNRNP70 (-4.03) |
| OGN (-4.99) | HSD11B2 (-5.38) | TPM4 (-3.72) |
| KRT86 (-4.42) | DCN (-5.36) | MYH10 (-3.61) |
| MRPL12 (-4.35) | CAVIN2 (-5.12) | TOP1 (-3.52) |
| GLIPR2 (-4.13) | CHGA (-5.04) | NEXN (-3.49) |
| NDUFS4 (-4.13) | HSPB6 (-5.04) | NCL (-3.46) |
| FABP1 (-4.10) | MYH11 (-4.86) | CALD1 (-3.41) |
| ZG16 (-4.07) | CKB (-4.59) | SUPT16H (-3.39) |
| ITLN1 (-4.06) | AOC3 (-4.50) | GUCY1B1 (-3.21) |

FC, fold change; NCG, Network of Cancer Genes, NM, normal mucosa; T, tumor; LM, liver metastasis

**Supplemental Table 8: Top 30 differentially expressed RNAs in two-group comparisons of patient 4.** Listed are the top 15 RNAs with increased or decreased expression in the comparison of tumor vs. normal tissue, metastasis vs. normal tissue, and metastasis vs. tumor tissue measured by log2FC. RNAs marked in bold are onco- or tumor suppressor genes annotated in NCG 7.0.

| T vs. NM | LM vs. NM | LM vs. T |
|---|---|---|
| **Top 15 RNAs with increased expression** | | |
| SERPINB5 (1.35) | LINC01819 (1.43) | POT1-AS1 (1.06) |
| CST1 (1.32) | APOA2 (1.41) | FGG (1.01) |
| MIR4500HG (1.15) | MIR4500HG (1.36) | LOC101926944 (0.98) |
| MIR4500 (1.14) | SERPINB5 (1.32) | MIR3144 (0.98) |
| KNG1 (1.14) | APOH (1.30) | ALB (0.95) |
| LHFPL3-AS2 (1.10) | MIR4500 (1.24) | MAP9 (0.92) |
| LINC01819 (1.09) | LHFPL3-AS2 (1.16) | SPP1 (0.89) |
| DNAH14 (1.05) | DEFA5 (1.11) | APOA2 (0.87) |
| CYP2B6 (1.03) | LGR5 (1.08) | GUCY1A2 (0.86) |
| PLEKHS1 (1.03) | GRIN2B (1.01) | MIR6888 (0.84) |
| ORM1 (1.03) | LINC02315 (1.01) | MIR603 (0.83) |
| DSG3 (0.99) | RNF128 (0.97) | RXFP1 (0.83) |
| ACE2 (0.98) | SAMD12-AS1 (0.97) | MIR5087 (0.81) |
| SAMD12-AS1 (0.98) | CYP2B6 (0.96) | PRG4 (0.81) |
| DMBT1 (0.96) | PLEKHS1 (0.96) | MIR8065 (0.81) |
| **Top 15 RNAs with decreased expression** | | |
| TMIGD1 (-1.43) | LIFR (-1.56) | PIGR (-1.35) |
| CR2 (-1.41) | CR2 (-1.55) | VIP (-1.04) |
| TLR10 (-1.39) | GCSAM (-1.53) | JCHAIN (-0.95) |
| VIT (-1.34) | CLCA4 (-1.50) | ACE2 (-0.90) |
| CLCA4 (-1.27) | FDCSP (-1.42) | DUOXA2 (-0.89) |
| GCSAM (-1.25) | CA1 (-1.40) | CNTN3 (-0.88) |
| LIFR (-1.24) | TLR10 (-1.40) | GZMK (-0.83) |
| MYBL1 (-1.23) | GZMK (-1.38) | GPR63 (-0.83) |
| CD22 (-1.20) | **AFF3** (-1.37) | KLRB1 (-0.82) |
| FDCSP (-1.20) | CR1L (-1.36) | **IL7R** (-0.80) |
| CD180 (-1.19) | LINC01215 (-1.32) | IGLL5 (-0.79) |
| **AFF3** (-1.19) | ANK2 (-1.32) | CD96 (-0.79) |
| OGN (-1.16) | TMIGD1 (-1.28) | CXCL14 (-0.78) |
| CR1L (-1.15) | CCDC152 (-1.26) | GPC5-AS1 (-0.77) |
| TTN (-1.15) | STAP1 (-1.24) | CRABP2 (-0.77) |

FC, fold change; NCG, Network of Cancer Genes, NM, normal mucosa; T, tumor; LM, liver metastasis
